# A Zero-Shot Decision Model (Jev) Matches a Trained Classifier but Ranks Its Own Errors Worse: Calibrated, Split-Conformal Next-Procedure Recommendation in General Dentistry

**DOI:** 10.64898/2026.05.07.26352635

**Authors:** Carson Conception Rodrigues, Steffie Dione Rebello

## Abstract

We evaluate calibrated, abstaining recommenders for what a dentist should do after the procedure just completed. On the public MEPS 2023 Dental Visits corpus (11,016 next-visit transitions, 5,088 patients, treatment-category granularity), a first-order history model reaches 0.479 top-1 (95% patient-clustered CI 0.457 to 0.500) against 0.297 for a most-frequent baseline, is well calibrated (expected calibration error, ECE, 0.031), and supports split-conformal selective prediction with a distribution-free coverage guarantee: on 1,637 held-out transitions, selective risk falls from 51.5% to 42.0% at 48.7% coverage. A zero-shot chat large language model (LLM) given the same history reaches 21.3% top-1 against 53.3% on a 150-transition probe. Jev, a decision model that returns a probability for every option, matches the trained model’s top-1 zero-shot (0.485; paired difference +0.008, CI −0.001 to +0.017) but is overconfident (ECE 0.098, 0.014 after isotonic recalibration) and ranks its own errors worse (area under the risk–coverage curve +0.077, CI +0.050 to +0.102), so the reject option is best built on the classical model. Because MEPS lacks procedure codes, we also built *DentaCoPilot*, a Current Dental Terminology-level recommender returning a top-*K* distribution, a confidence label, an abstain flag and a rationale, and compared classical and LLM designs on a synthetic corpus as a pipeline check. On 200 stratified examples Sonnet reaches 0.360 against 0.500 for the classical model (*p*=0.0003), and 0.440 with the classical model’s candidates in its prompt, still below it (*p*=0.036); Jev Router, given the same prompts, reaches 0.300 and 0.425, matching Sonnet with the prior (*p*=0.66). Validation on real charts and a clinician-in-the-loop study remain future work.

## 1. Introduction

Commercial dental AI in 2026 is overwhelmingly diagnostic. The most recent FDA-clearance survey [1] catalogues 29 cleared products from 13 companies (Overjet, Pearl, VideaHealth, Dexis, Perimetrics, and others) targeting single-image classification problems: caries detection, calculus identification, periapical radiolucency, and periodontal bone-level measurement.^2^ Independent academic evaluations of these products (of Overjet’s caries module [3] and Pearl’s Second Opinion [4]) confirm their diagnostic-only scope, and an umbrella review of caries-detection AI [5] (137 studies; pooled sensitivity 0.85, specificity 0.90) concludes such tools should augment rather than replace clinical judgement. Peer-reviewed evidence for clinical benefit exists for the caries-detection variant in particular: in a cluster-randomised cross-over trial of 22 dentists assessing bitewings, Mertens et al. [6] reported that AI-assisted review raised the mean diagnostic-accuracy area under the ROC from 0.85 to 0.89 (ΔAUC= + 0.04, *p<*0.05); a follow-up trial confirmed the gain was cost-effective at practice scale [7]. These are valuable deployments. They are also, individually, single-decision tools. They do not answer the clinically harder question that follows every diagnosis: *what should the dentist do next?*

Sequencing a treatment plan from a chart is a different problem. A two-surface posterior composite on tooth 19 leads to one set of likely continuations; the same composite on a tooth with a prior endodontic history leads to another; in a periodontitis patient with diabetes, neither sequence is appropriate. The closest published system that operates at this layer, MultiTP [8], is a CNN-RNN restricted to partial-edentulism cases with a fixed five-treatment output sequence. It does not surface calibrated uncertainty, does not abstain when the chart context is insufficient, and was not evaluated as a clinician-augmenting tool. The largest related effort in periodontitis-risk prediction [9] is a risk model, not a procedure recommender. A concurrent multi-site effort, RT2C [10], predicts time-to-new-caries with a recurrent network over 466,782 patients but targets a single future event rather than the next procedure; and an analysis of restoration-survival prediction on the BigMouth repository [11] finds that dental electronic records are not yet uniformly ready for accurate procedure-outcome prediction, which motivates careful task scoping and our augmentation framing.

We propose *DentaCoPilot*: an LLM-augmented next-procedure recommender designed from the start as decision support for dentists. The system takes a structured chart and the most recently completed procedure, and emits four signals together: (i) a top-*K* probability distribution over Current Dental Terminology codes for the next procedure; (ii) verbalised confidence in {low, medium, high}; (iii) an explicit abstain flag when the chart context is genuinely insufficient; (iv) a chart-grounded rationale citing specific prior procedures and findings. The model is constrained-decoded to the CDT 2024 vocabulary and calibrated via temperature scaling. The contribution lies less in raw accuracy than in the framing: we evaluate the system as a consult that augments the dentist, not as a classifier that replaces the dentist’s judgement.

We test two hypotheses on a synthetic chart corpus of 500 patients (§3.2): **(H1)** a calibrated LLM-based recommender can match classical-baseline accuracy on next-procedure prediction while preserving rationale and abstention; **(H2)** injecting a classical baseline’s top-*K* candidates into the LLM prompt is the most cost-effective LLM design for achieving (H1). We evaluate four classical baselines, six LLM variants spanning Claude Haiku, Sonnet, and Opus with chain-of-thought, retrieval, and prior-conditioning, and report calibration and coverage–risk analyses. We then test external validity for the task on real-world data using the public MEPS 2023 Dental Visits corpus [12] (§4.9), at treatment-category granularity. A prospective clinician-in-the-loop evaluation, and CDT-level multi-institutional validation [13, 14] (gated behind institutional data-use agreements), remain future work, contingent on clinical-data access and ethics approval that we have sought but not secured. Every CDT-level result in this paper is measured on synthetic charts; the only real-data evidence is at treatment-category granularity (MEPS), and CDT-level validation on real charts is left to future work.

The paper’s main empirical result is therefore on real data: on MEPS, a calibrated history-aware classical model outperforms a zero-shot chat LLM given the same category history, is well calibrated (expected calibration error 0.031), and supports split-conformal abstention with a distribution-free coverage guarantee. The synthetic CDT-level comparison of classical and LLM designs is reported as a pipeline check, with paired tests and intervals at *n*=30 and in a category-stratified *n*=200 re-run, and the LLM’s chart-grounded value is located in the chart-rich regime that neither corpus can yet test on real data.

We also evaluate TypeSafe’s Jev [15], a decision model that returns a probability for every option instead of text, and Jev Router [16], which sends each chat request to an underlying LLM of its choosing. Because Jev returns a full probability vector, it is the one model from the LLM side of this study that enters the calibration and conformal analysis on real MEPS data on the same terms as the classical model. There, zero-shot, it matches the trained model’s accuracy but is overconfident, and even after recalibration it ranks its own errors worse, so the reject option is still best built on the classical model (§4.10).

All LLM inference in this work is routed through the local Anthropic Claude Code CLI [17], with full per-call audit logs released alongside the code (see §3.6). The Jev runs are the exception and use the OpenRouter API, with the model that served each request logged.

## 2. Related Work

### Diagnostic dental AI in commercial deployment

Shujaat [1] catalogues the FDA-cleared products to date: caries / calculus detection (Overjet, Pearl, VideaHealth), periapical radiolucency (Overjet, Pearl), automated dental charting (Overjet), millimetre-precision bone-level quantification (Overjet, Pearl), 3D cone-beam CT segmentation (Pearl), and acoustic fracture detection (Perimetrics). These systems share a common shape: an image goes in, a finding comes out. None plans treatment.

### Procedure sequencing

MultiTP [8] is the closest published system to what we propose: a CNN-RNN with attention that predicts up to five sequential treatments for partial-edentulism cases, achieving an AUC of 0.91 over five plans. It is an EDR-only model with no calibration, no abstention, and no clinician-augmentation evaluation. We reimplement a minimal MultiTP-style architecture as our B3 comparator. In a related specialty domain, Li and Wang [18] use multi-task reinforcement learning with explainable AI to plan orthodontic-orthognathic treatment, but the task is sufficiently different (continuous decision-making over a longer planning horizon, narrow specialty) that the contributions do not transfer directly to general dentistry.

### Risk prediction in dentistry

Patel et al. [9] model periodontitis risk on 20,946 adult patients with linked EHR-EDR data, demonstrating that procedure-level structured data can support clinical prediction; the same group has also shown that adding social determinants of health to such models meaningfully changes risk calibration [19]. Varghese et al. [20] pair Cariogram-based caries-risk profiling with parent-oriented educational mobile messages in a randomised trial, an example of coupling structured risk modelling with a patient-facing digital intervention. All three are risk models rather than procedure recommenders; we position our work as the procedure-recommendation analogue.

### LLMs in dentistry

Farhadi Nia et al. [21] surveys early LLM applications in dental practice, focusing on documentation, summarisation, and patient-facing chat. The review explicitly flags that LLMs “present clean, fluent statements without communicating uncertainty,” which is the failure mode this paper directly addresses through calibration and abstention. Broader medicine-LLM work [22, 23] shows that capable models reach physician-level accuracy on knowledge benchmarks, but Omiye et al. [24] demonstrates that the same models can propagate dangerous race-based clinical claims, which motivates our hard requirement for chart-grounded rationale and explicit abstention. Schwendicke et al. [25] surveys the chances and challenges of dental AI broadly; recent systematic and scoping reviews of LLMs in dentistry [26, 27] map a literature concentrated on diagnosis, education, and patient communication, and a dental-specific position statement on generative AI [28] stresses verification and human oversight, directly motivating our chart-grounded-rationale and abstain design. The nearest CDT-level LLM work extracts codes from clinical notes rather than forecasting the next one, and reports plausible but wrong codes [29]. Based on a structured search through October 2026, we are not aware of prior work that builds and evaluates an LLM as a treatment-*sequence* recommender under a clinician-augmentation (rather than autonomy) framing.

### LLMs versus classical models on structured clinical prediction

A cross-domain literature has begun to ask whether general-purpose LLMs beat established machine-learning models on structured clinical prediction, and the answer so far is largely negative. ClinicalBench [30] benchmarks 22 general-purpose and medical LLMs against 11 traditional models and reports that the LLMs do not yet beat methods such as SVM and XGBoost; the count-based EHR study of Gao et al. [31] similarly finds that gradient-boosted and TabPFN learners remain competitive with transformer and LLM pipelines on structured records, and ClinicRealm [32] re-evaluates the comparison across many tasks and reaches a mixed verdict in which conventional models hold their own; Ko et al. [33] report the opposite for ASA classification from structured EHR data, and outside dentistry Rajesh et al. [34] pair LLM next-treatment recommendations with uncertainty-based de-ferral. Our results replicate this pattern in the dental next-procedure setting for chat LLMs: a zero-shot chat LLM trails the classical baseline on real category-history data, and the pure-LLM variants trail the classical baselines on the synthetic corpus, with paired contrasts that are significant for Haiku at *n*=30 and for Sonnet in a stratified *n*=200 re-run (§4.5, §4.7). Our contribution is to show that prompt-conditioning a small LLM on the classical baseline’s top-*K* candidates (M5) then recovers most of that gap while retaining the calibration, abstention, and chart-grounded-rationale layer that an autonomous classifier does not provide. A decision model that returns probabilities qualifies the pattern: Jev matches the classical model’s accuracy on MEPS zero-shot, and the classical model’s advantage moves to calibration and to how well its confidence ranks its own errors (§4.10).

### Calibration and abstention in clinical AI

Our calibration approach follows Guo et al. [35] (temperature scaling) with Platt [36] as the precursor. Coverage– risk and selective prediction are standard tools in safety-critical ML; we apply them to the procedure-recommendation setting.

### Datasets

BigMouth [13, 14] is a multi-institutional dental EHR repository with 6M+ patients across multiple dental schools, accessed under a project-review-committee proposal. Open imaging datasets relevant for chart- context grounding include DENTEX [37], MMDental [38], and DenPAR [39].

## 3. Methods

### 3.1. Task formulation

We formulate next-procedure prediction as a single-step classification over the Current Dental Terminology code set [2]. A patient chart is a tuple *C* = (*D, M, H*) where *D* holds demographics (age band, sex), *M* is a list of medical-history flags, and *H* = (*e*_1_*, e*_2_*, . . ., e_T_*) is a time-ordered sequence of procedure events. Each event *e_t_* = (*τ_t_, c_t_, k_t_*) carries a date *τ_t_*, a CDT code *c_t_* ∈ V, and an optional Universal-numbered tooth identifier *k_t_* ∈ {1*, . . .,* 32} ∪ {⊥} in the Universal Numbering System [40].

Given a chart truncated at index *t* (i.e. the dentist has just performed *e_t_*), the model outputs:

1. A top-*K* probability distribution *p*^(*c_t_*_+1_ | *C*_1:_*_t_*) ∈ Δ^|V|^ over the next procedure code, returned as the top-*K* codes ranked by probability.
2. A confidence label *q*^ ∈ {low, medium, high}, a verbalised self-rating [41, 42, 43, 44].
3. An abstain flag *a* ∈ {0, 1} which the system sets to 1 when the chart context is judged insufficient.
4. A short rationale *r* grounded in cited fields of the chart *C*_1:_*_t_* (used for evaluation, not for the prediction itself).

We expand each chart of length *L* into *L* − 1 prediction examples by truncating at every prefix index *t* ∈ {1*, . . ., L* − 1}. Train, dev, and test splits are constructed at the *patient* level (no chart appears in two splits), following standard EHR-modeling practice [45, 46].

We report **top-***K* **accuracy**: the fraction of test examples for which the gold next-procedure code appears among the model’s *K* highest-probability predictions. Top-1 corresponds to exact CDT match; top-3 and top-5 capture clinically reasonable shortlists for a recommender (a dentist seeing a small candidate list with calibrated probabilities is the deployment mode we have in mind). We also report top-1 by CDT category and, where calibrated probabilities are available, Expected Calibration Error and coverage–risk under abstention (§3.4).

### 3.2. Datasets

*Synthetic chart corpus..* Because CDT-coded real-data access was sought but not secured, the *CDT-level* experiments use a synthetic corpus generated by a hierarchical Markov-style procedure-history generator; real-world external validity at category granularity is provided separately by MEPS (below). Because this generator is a Markov/bigram process, accuracy comparisons among models that themselves encode bigram structure (B0/B1 in particular) are partly circular; we therefore treat the synthetic arm as a test of pipeline behaviour and of the augmentation properties (rationale, calibration, abstention), and reserve externally valid accuracy claims for real data. Patients are assigned to one of five trajectory archetypes (*healthy*, *restorative-prone*, *periodontitis-prone*, *edentulous-progressing*, *ortho-adolescent*) with plausibility-based priors. Within each archetype, per-archetype transition tables encode bigram probabilities Pr(*c_t_*_+1_ | *c_t_,* archetype) drawn from clinical workflow knowledge and the published procedure-frequency analyses in the BigMouth corpus [13, 14]. Tooth-level state is tracked: extracted teeth cannot receive subsequent restorations; root-canal-treated teeth tend to receive crowns next; SDF-treated sites revert to monitoring [47, 48]. We generate 500 patients over a five-year timeline, split 70/15/15 at the patient level (350 / 75 / 75 charts), and expand to 8,427 prediction examples (5,878 train / 1,265 dev / 1,284 test). As a face-validity check, we compared the generator’s visit-level category dynamics against the real MEPS corpus under a common nine-category scheme. Agreement is *weak*: marginal next-category distributions correlate at only Pearson *r* = 0.19 (total-variation distance 0.46) and first-order transition probabilities at *r* = 0.50. The generator over-represents standalone examinations and under-represents routine cleanings/prophylaxis and prosthodontic care relative to the US visit mix, reflecting its clinical-archetype design and the population/granularity gap. We therefore read the synthetic accuracy results strictly as tests of pipeline behaviour and of the augmentation properties, not as estimates of real-world performance; externally valid accuracy is contingent on the real-data (MEPS, below) and on future real-data evaluations (meps_synth_fidelity.py).

#### Public imaging datasets (planned, for chart-context grounding)

Loaders are implemented and verified for DENTEX (MICCAI 2023 grand-challenge [37]; 2,332 panoramic X-rays with caries, deep caries, periapical, and impacted-tooth labels); MMDental [38], with 660 paired CBCT volumes and structured records; and DenPAR [39], with annotated periapical radiographs. We do not auto-download these datasets; the loaders document the manual fetch step and verify SHA-256.

#### Real-world public benchmark (MEPS)

To complement the synthetic corpus with real-world data requiring no controlled-access agreement, we use the Medical Expenditure Panel Survey 2023 Dental Visits public-use file [12] (AHRQ HC-248B; 19,599 dental events for 8,583 persons). MEPS records, per visit, eleven treatment-type indicators (examination, cleaning/prophylaxis, preventive [fluoride/sealant], restorative, endodontic, periodontal, oral surgery, implant, prosthodontic, orthodontic, and other) but *not* CDT codes; we therefore evaluate at *category* granularity. We assign each visit its single most clinically significant procedure by a fixed priority (implant *>* oral surgery *>* endodontic *>* periodontal *>* prosthodontic *>* orthodontic *>* restorative *>* preventive *>* prophylaxis *>* examination), order each person’s visits by date, and form next-visit transitions, yielding 11,016 transitions over 5,088 patients with ≥ 2 visits. The task mirrors the synthetic task one granularity coarser: given the most recent visit’s category (and provider type), predict the next visit’s category. All MEPS analyses use raw next-visit transitions and do *not* apply the MEPS survey weights, so every estimate describes the sampled cohort rather than nationally weighted US totals.

#### Real CDT-level data (future work)

Controlled access to the BigMouth Dental Data Repository [13, 14] was sought but not granted; CDT-coded multi-institutional repositories remain gated behind US-institution data-use agreements and are named as future work. A retrospective chart pull at a clinical partner would provide single-institution real CDT-level data and underpin a prospective clinician-in-the-loop evaluation; any such study remains contingent on clinical-data access and ethics approval that we have sought but not secured.

### 3.3. Models

We evaluate four classical baselines and six LLM variants. All operate on the same patient-level splits and the same prediction examples. CDT codes outside the training-set vocabulary are masked out at inference; LLM responses are post-hoc constrained to the training vocabulary.

#### Classical baselines

**B0 frequency bigram** *p̂*(*c_t_*_+1_ | *c_t_*) computed from training data with add-one smoothing. The trivial sanity floor.
**B1 TF-IDF + multinomial logistic regression** [49, 50]. Tokens include each historical CDT code, positional-from-end markers for the last three procedures, and categorical demographics. Library defaults, untuned (*C*=1.0).
**B2 XGBoost** [51] on engineered chart-history features: counts per CDT category, has-extraction / has-RCT / has-implant / has-crown booleans, time since last visit, age band one-hot, sex, and a small medical-history flag set. 200 trees, depth 6, learning rate 0.1, set a priori and untuned.
**B3 MultiTP-style CNN–RNN** [8]. Minimal reimplementation: 32-dim code embedding, 1-D CNN over the most recent procedures (analogous to MultiTP’s tooth + neighbour tensor), GRU over the longer sequence, attention pooling, softmax head over the CDT vocabulary. ∼30k parameters, 12 epochs.

#### LLM variants

All LLM inference is routed through the local Anthropic Claude Code CLI (§3.6), not the metered Anthropic API.

**M1 Claude Haiku** (`claude-haiku-4-5-20251001`). Structured prompt: chart pretty-printed, top-*K* instruction, JSON-only output schema, top-40 most-frequent training codes shown as a vocabulary palette.
**M2 Claude Sonnet + chain-of-thought** (`claude-sonnet-4-6`). Same prompt as M1 but with an explicit <thinking>…</thinking> reasoning step before the JSON, in line with prior work showing CoT prompting improves reasoning-heavy clinical tasks [52, 23].
**M3 Claude Sonnet + retrieval** (*M* 2 + BM25 [53]). The top-3 BM25-retrieved cards from a twenty-card open-source clinical evidence corpus (paraphrased from AAE / AAP / AAOMS standard practice; original ADA-copyrighted material is not reproduced) are injected into the prompt; the model is instructed to cite specific cards (`[E###]`) in its rationale.
**M4 Claude Opus + chain-of-thought** (`claude-opus-4-7`). Identical prompt and schema to M2; only the model identifier changes. Tests whether more capable backbone helps.
**M5 Sonnet + classical prior** *The proposed forward design*. The B1 (TF-IDF + LR) baseline is fit on the same training set; its top-10 (code, probability) candidates are injected as a prior into the M2 prompt. The LLM is instructed to re-rank, redistribute, or override.
**M6 Opus + classical prior** M5 with Opus backbone. Tests whether priming compounds with capability.

#### Jev designs (added October 2026)

TypeSafe’s Jev [15] is a *decision* model rather than a chat model: given a structured state and a choice question with named options, it returns the chosen option and a probability for every option, and its developer describes it as trained for calibrated decisions. Jev Router [16] uses Jev to send each chat request to an underlying LLM and reasoning effort chosen for quality, speed and cost; learned routing across medical LLMs has been proposed independently [54]. Both are reached only through OpenRouter, so these runs bypass the Claude Code wrapper (§3.6). We add four designs, each paired example-for-example with an existing run.

**M7 Jev Router + chain-of-thought** (`typesafe/jev-router`). The M2 prompt, byte-identical (checked against all 200 logged Sonnet prompts before any call), on the stratified *n*=200 sample of §4.7.
**M8 Jev Router + classical prior** The M5 prompt, byte-identical, on the same 200 examples.
**J0 Jev zero-shot (MEPS)** (`typesafe/jev-1.13`, snapshot 20260917). One eleven-option choice question per MEPS transition, with option descriptions taken from the MEPS treatment flags and the category priority rule stated; the state is the most recent visit’s category and provider type, the same two features the MEPS logistic baseline uses. No training data are shown.
**J1 Jev + training frequencies (MEPS)** J0 with the state extended by the next-visit category frequencies that followed the same history in the MEPS *training* split, and their count: the decision-model analogue of M5’s prompt-conditioning.

Because J0 and J1 see only two categorical features, their output is a function of the 44 distinct (category, provider) states; we queried each state five times per design (440 calls), used the first answer per state as the primary analysis (what a single-call deployment sees), and report the mean over the five as a sensitivity analysis and the spread as a stability measure. Jev Router was also run on the 150-transition MEPS probe of §4.9 with the Haiku prompt unchanged.

### 3.4. Calibration and abstention

We apply temperature scaling [35, 36] to the top-1 probability of each model. The temperature *T* is fit on a random half of the test split by minimising negative log-likelihood [55]; the rescaled top-1 probability is reported on the held-out half. We compute the Expected Calibration Error [55, 56] on equal-width bins.

For abstention, we sweep a threshold *τ* over the top-1 probability and report coverage–risk curves [57, 58]: over the temperature-scaled probability for the coverage-precision table of §4.3, and over the raw B1 probability for the selective-prediction and out-of-distribution analysis of §4.4. On the synthetic corpus we report a single illustrative operating point, with *τ* set on the test split to retain 80% coverage; this is an in-distribution demonstration, not a safety threshold, and it carries no coverage guarantee. The principled, guarantee-carrying selective-prediction rule is the split-conformal reject option [59] that we apply to the real MEPS data (§4.9), whose accept threshold is calibrated on a held-out split and whose coverage is distribution-free.

### 3.5. Hybrid integration

We test two ways to combine classical and LLM predictions:

1. **Confidence-routed gating.** Pick the baseline’s output when its top-1 probability ≥ *τ* ; otherwise the LLM’s. We sweep *τ* ∈ {0, 0.2*, . . .,* 1.0}.
2. **Prompt-conditioning (M5/M6).** The classical model’s top-10 (code, probability) candidates are part of the LLM prompt; the LLM produces a single output that uses, refines, or overrides the prior.

(2) is implemented end-to-end as M5 and M6; (1) is computed post-hoc from the predictions logged by each model.

### 3.6. Compute infrastructure

Every LLM inference call in this paper runs through a single subprocess wrapper at `data/cc_compute.py`, which invokes the local Anthropic Claude Code CLI [17] with `--output-format json` and parses the response. The wrapper logs every call (timestamp, run id, model, prompt, response, latency, input and output token counts) to a per-run `llm_calls.jsonl`, which the repository commits as a permanent record. We deliberately do not import the Anthropic Python SDK; a continuous-integration grep guard (`check-no-anthropic` in the Makefile) prevents accidental introduction. This design (a) routes compute to the existing Claude Code subscription, (b) provides, for the synthetic-corpus runs, a complete audit trail every reviewer can re-read without spending compute, and (c) makes the experiment reproducible by simple replay instead of re-execution.

The Jev designs are the one exception: they run through `data/openrouter_compute.py`(Jev Router) and `data/meps_jev_decisions.py` (Jev), which call the OpenRouter API and log the same fields plus what a router makes necessary: the model that actually served each request, the upstream provider, the generation identifier and the billed cost. A router may send identical prompts to different models, and its routing policy and model pool are not published and may change [16], so the Jev Router results can be audited from the logs but not re-executed exactly. All Jev calls were made on 10 October 2026 and cost US$1.87 in total; they carried only synthetic charts and public MEPS category states.

### 3.7. Reproducibility protocol

We structure model development and reporting to follow the TRIPOD+AI [46] and TRIPOD-LLM [60] reporting guidelines (completed checklists in Supplementary Material S1); any future prospective clinician-in-the-loop study (§4.9) would also follow DECIDE-AI [61] for early-stage clinical evaluation, and imaging-specific reporting (CLAIM) applies only to the future imaging-grounding arm.

All seeds are fixed (`random.seed`(42), `numpy.random.seed`(42), `torch.manual_seed`(42)). Each experimental run writes `config.json` (git SHA, seed, CLI args, Python and platform version), `results.json` (per-trial predictions and gold labels), and `summary.json` (aggregated metrics). The synthetic generator, all baseline and LLM implementations, the evaluation harness, and the analysis scripts are archived at Zenodo (doi:10.5281/zenodo.21059895) under the MIT licence (see the Data and code availability statement).

## 4. Experiments

### 4.1. Classical baseline accuracy at scale

We first evaluate the four classical baselines (B0–B3, defined in §3.3) on the full test split (1,284 examples). This is the larger-*n* regime; LLM evaluation in §4.5 is necessarily smaller.

**Table 1:**
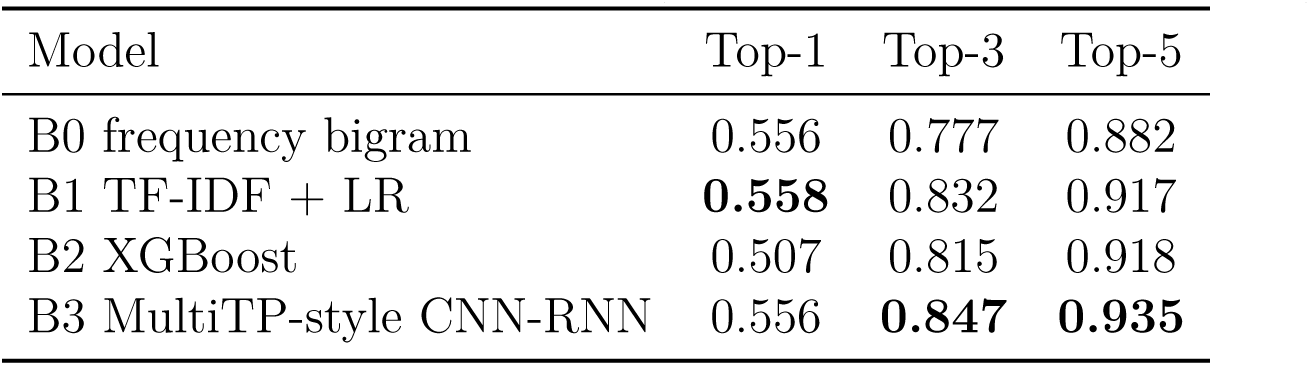
Synthetic-corpus baseline results (*N* =500 charts; 1,284 test examples).

The four baselines cluster tightly on top-1 (50.7–55.8%) but spread on top-K>1. The MultiTP-style sequence model (B3) wins both top-3 (0.847) and top-5 (0.935), suggesting that CNN+RNN attention captures useful ordering of treatment continuations beyond what a flat bigram or feature classifier can. Top-1 is harder: the bigram baseline’s strong local Markov signal nearly ties B1 (0.556 vs 0.558), and XGBoost lags both, indicating the engineered feature set under-uses recent-procedure information.

The per-category breakdown (Figure 2) reveals the opportunity for chart-aware reasoning. On the full test split (*n*=1,284), classical baselines reach 56–80% top-1 on bundled / sequential procedures (diagnostic 65–75%, adjunctive 67–80%, orthodontic 62–78%, preventive 56–59%) but *collapse* on the multi-class restorative choice (0–7%), endodontic sequencing (0%, scarce in synthetic data), and removable prosthodontic decisions (0%). This is exactly the regime where a chart-aware LLM with calibrated abstention should help. The next subsection (§4.5) reports the LLM evaluation against these baselines.

**Figure 1:**
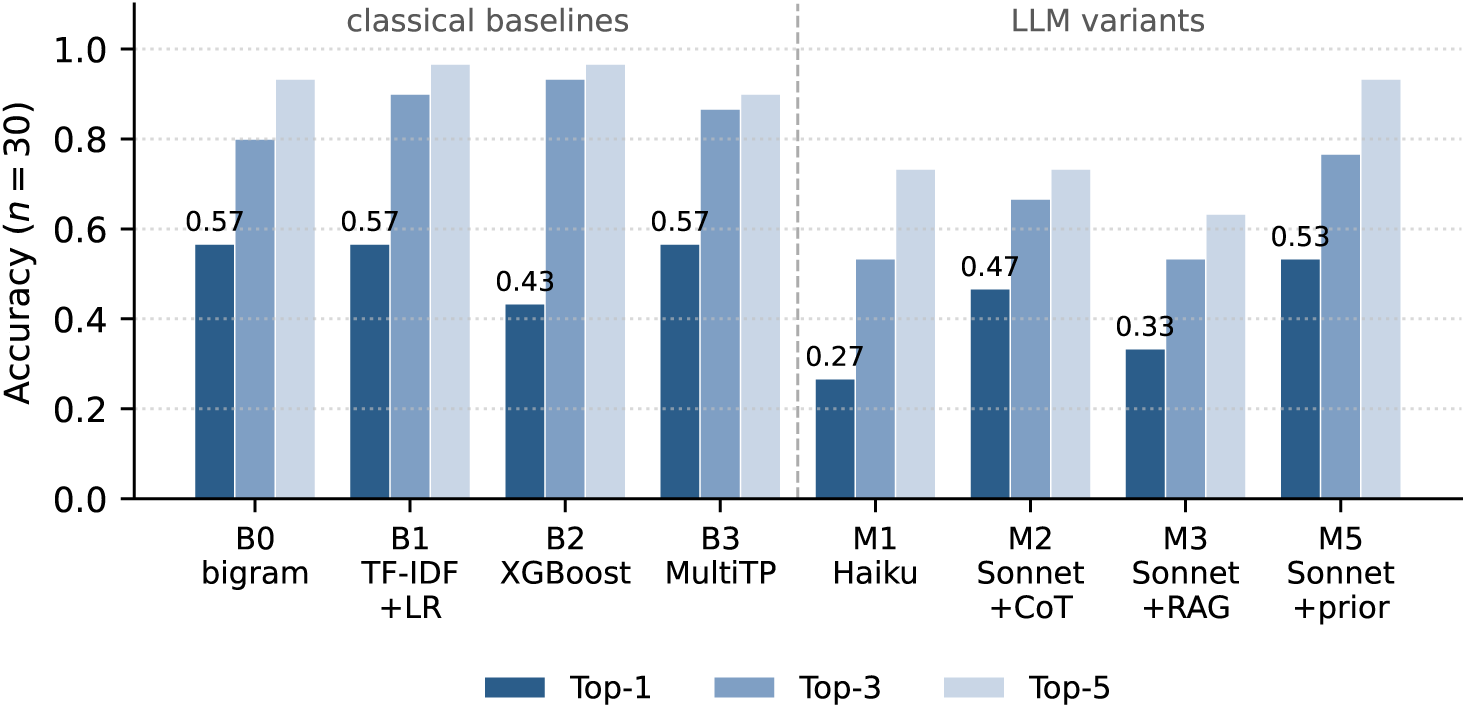
Top-*K* next-procedure accuracy on the matched *n*=30 synthetic subsample (top-1 values labelled). Three classical baselines (B0, B1, B3) reach 0.567 top-1, above every pure LLM variant (M1–M3, 0.267 to 0.467); B2 XGBoost (0.433) is below M2 on top-1 but leads on top-3 and top-5. Prompt-conditioning Sonnet on the classical candidates (M5) reaches 0.533 top-1 and 0.933 top-5. At this sample size only Haiku’s gap is significant; the stratified *n*=200 re-run tests the contrasts with power.

**Figure 2:**
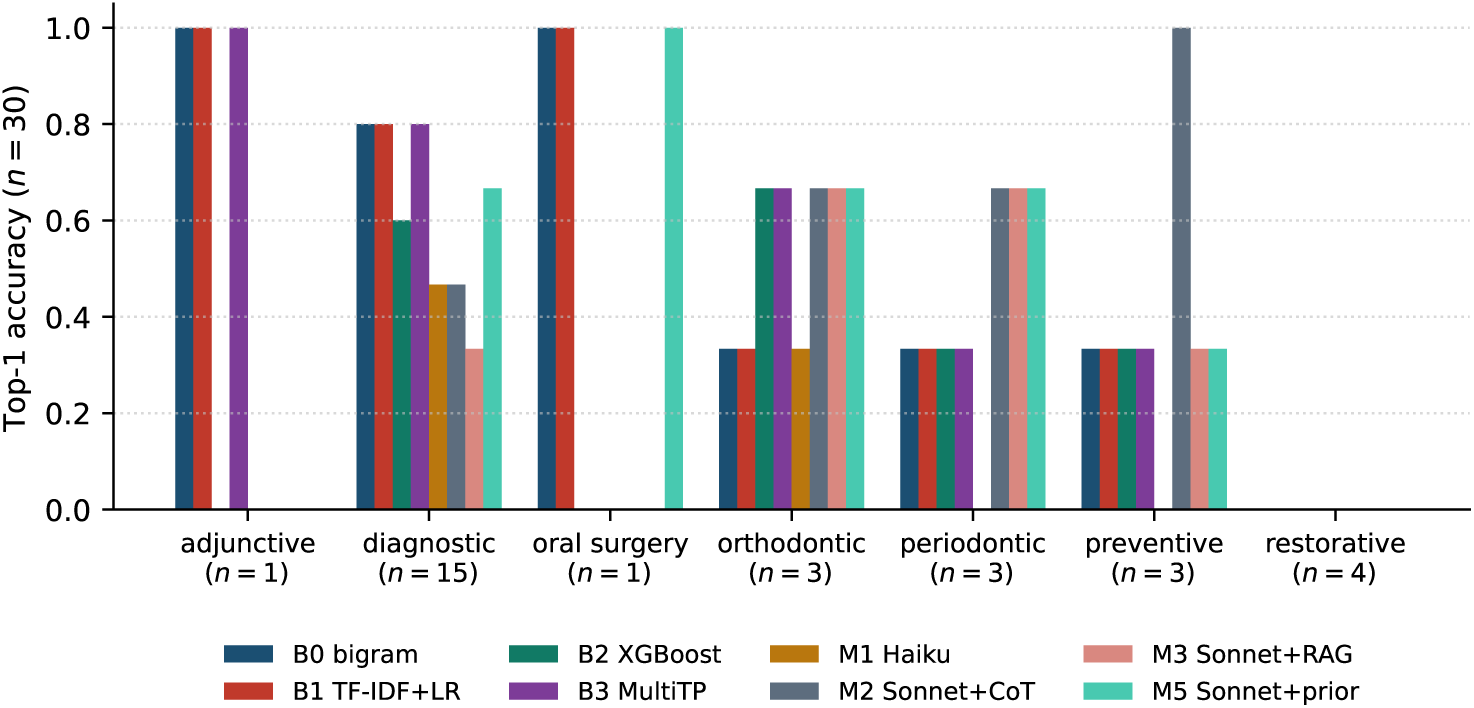
Top-1 accuracy by CDT category on the matched *n*=30 subsample (category sizes under the labels). Classical baselines lead on diagnostic visits (0.60–0.80 against at most 0.67 for any LLM design); the single adjunctive case is solved only by B0, B1 and B3, and the single oral-surgery case by B0, B1 and M5. The Sonnet designs (M2, M3, M5) reach 0.67 on periodontic cases against 0.33 for every classical baseline, and M2 is 1.0 on preventive cases against 0.33. Most cells hold one to four examples and are illustrative only. No model gets a restorative case right: the codes differ only in surface count.

### 4.2. Reproducibility (run records)

Every experimental run is recorded as described in §3.7. Per-run `config.json, results.json, summary.json`, and (for LLM-backed runs) llm_calls.jsonl are included in the public deposit at Zenodo (doi:10.5281/zenodo.21059895) (see the Data and code availability statement). All large language model inference in this paper runs through the local Anthropic Claude Code CLI instead of the metered Anthropic API; the wrapper at `data/cc_compute.py` is the single chokepoint.

### 4.3. Calibration and abstention

Following the protocol in §3.4, we post-hoc calibrate each baseline and report ECE pre- and post-temperature-scaling.

**Table 2:**
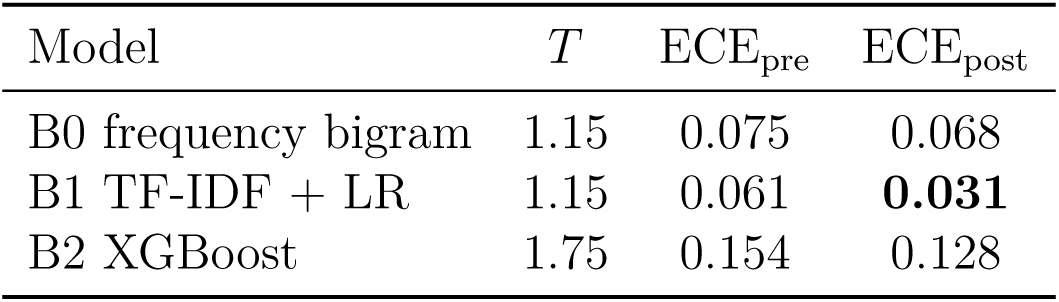
Temperature scaling and ECE. *T* is fit on a random half of the 1,284 synthetic-corpus test examples; both ECE columns are computed on the other, held-out half (*n*=642), ten equal-width bins.

| Model | $T$ | $\text{ECE}_{\text{pre}}$ | $\text{ECE}_{\text{post}}$ |
| --- | --- | --- | --- |
| B0 frequency bigram | 1.15 | 0.075 | 0.068 |
| B1 TF-IDF + LR | 1.15 | 0.061 | **0.031** |
| B2 XGBoost | 1.75 | 0.154 | 0.128 |

XGBoost is materially overconfident (*T* =1.75) and remains the worst calibrated model after scaling. The bigram and TF-IDF + LR models are already reasonably calibrated and improve modestly; with 642 examples and ten bins, ECE has a noise floor of roughly 0.02, so differences of that size between models should not be over-read, and the rescaling does not change the ranking among models.

Applying the abstention rule “cover only when the calibrated top-1 probability ≥ *τ* =0.6” yields the coverage-precision trade-off in Table 3:

**Figure 3:**
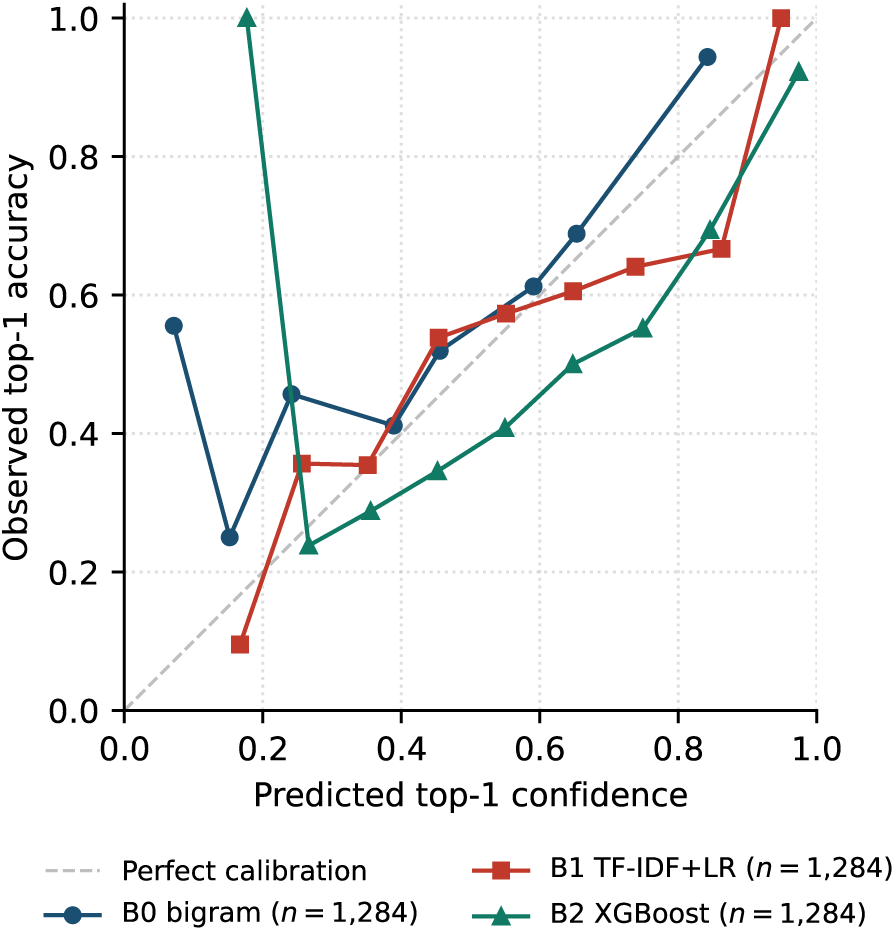
Reliability diagram for the three primary baseline models on the full *n*=1,284 baseline test split (LLMs omitted because *n*=30 is too small for stable per-bin estimates). The dashed diagonal is perfect calibration. B2 XGBoost is materially overconfident in the mid-range; B0 and B1 hew closer to the diagonal.

**Table 3:** Coverage and precision under abstention at *τ* =0.6 on the temperature-scaled top-1 probability (all 1,284 test examples).

| Model | Coverage | Top-1 (overall) | Top-1 (covered) |
| --- | --- | --- | --- |
| B0 frequency bigram | 0.21 | 0.556 | **0.772** |
| B1 TF-IDF + LR | 0.34 | 0.558 | 0.731 |
| B2 XGBoost | 0.39 | 0.507 | 0.706 |

The bigram baseline trades coverage for precision aggressively: it abstains on 79% of cases but answers correctly 77% of the time when it does. This is the failure mode the LLM is expected to *mitigate*: matching that precision while abstaining on far fewer cases.

### 4.4. Selective prediction and out-of-distribution detection

The verbalised `abstain` flag each LLM emits never fired on the synthetic test set (§4.5), so it cannot serve as the system’s safety control. We therefore build abstention on the classical model’s top-1 probability rather than on the model’s own self-report. For this analysis the score is the uncalibrated top-1 probability of the B1 model, with *τ* set to the 20th percentile of the test-split scores (*τ* =0.36) so that coverage is 80% by construction; the operating point is chosen on the test split and is an in-distribution demonstration, not a safety threshold (§3.4).

At that operating point (Table 5), abstaining on the lowest-confidence fifth of cases lowers the error rate among covered predictions from 44.2% (answer everything) to 38.1%: a 6.1-point reduction in risk on the cases the system chooses to act on. The curve is monotone, so a deployment willing to abstain more can trade coverage for safety (dropping to 50% coverage pushes covered risk down to 31.6%).

**Table 4:** Split-conformal selective prediction on the real MEPS test set (history-aware logistic model; calibration and evaluation are disjoint patient halves, *n*_eval_=1,637 transitions / 764 patients). Achieved coverage tracks the conformal target within sampling error (bootstrap 95% CI over patients), and selective risk (error among accepted) decreases monotonically as coverage tightens. ECE on the evaluation half is 0.031.

| Target cov. | Achieved cov. | 95% CI | Selective risk |
| --- | --- | --- | --- |
| Answer everything | 1.000 | n/a | 0.515 |
| 0.90 | 0.878 | [0.856, 0.898] | 0.491 |
| 0.80 | 0.798 | [0.773, 0.822] | 0.474 |
| 0.70 | 0.713 | [0.683, 0.742] | 0.450 |
| 0.60 | 0.645 | [0.612, 0.676] | 0.444 |
| 0.50 | 0.487 | [0.454, 0.524] | **0.420** |

**Table 5:**
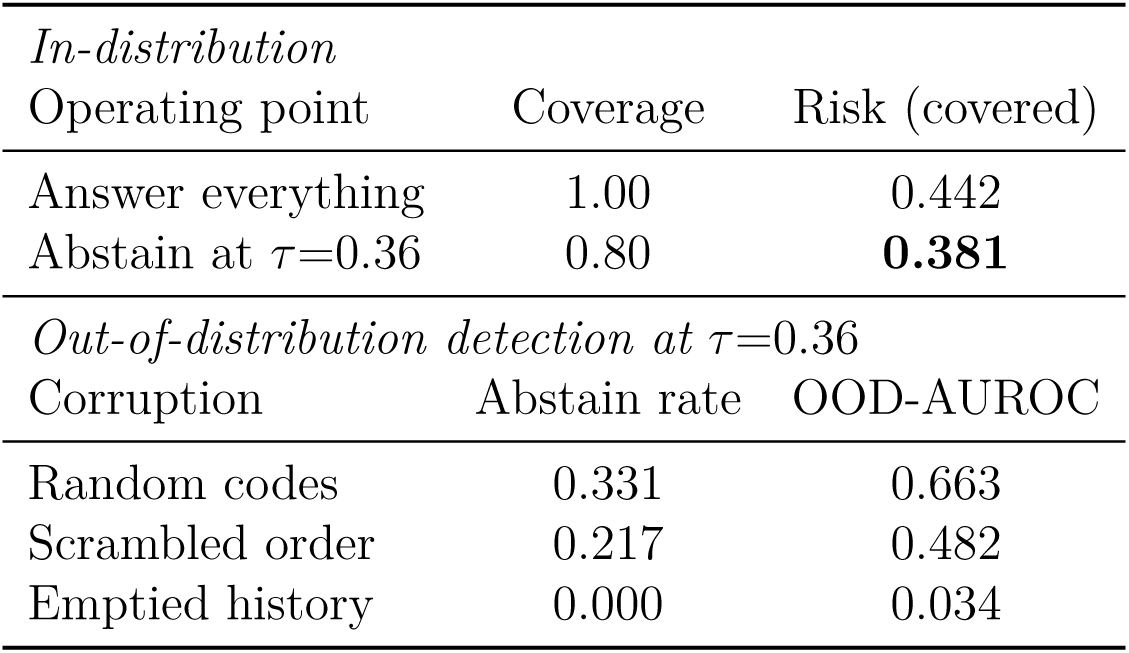
Selective prediction on the raw (uncalibrated) B1 top-1 probability (*n*=1,284 test examples; *τ* is the 20th percentile of these scores). *In-distribution*: error among covered cases at the 80%-coverage operating point versus answering every case. *Out-of-distribution*: abstention rate and OOD-detection AUROC for three corruption families at the same threshold (*τ* =0.36).

We then test whether the same threshold flags inputs the model should not answer at all. We build three out-of-distribution chart families from the test set and, at the fixed *τ*, measure how often the rule abstains and how well the calibrated probability separates in- from out-of-distribution inputs (area under the ROC, OOD-AUROC). On charts with randomly substituted procedure codes the rule abstains 33.1% of the time (AUROC 0.663): a usable but modest detector. On order-scrambled histories it abstains only 21.7% (AUROC 0.482, no better than chance), and on emptied single-event histories it abstains 0% (AUROC 0.034), failing exactly where the chart context is thinnest. Probability-threshold abstention therefore buys a real in-distribution risk reduction but is not by itself a sufficient out-of-distribution safeguard; principled selective-prediction methods, learning-to-defer [62] and the conformal reject-option rule we apply to the real MEPS data below, are the appropriate next step. The OOD-construction code and full results are released (`abstention_ood.py`).

**Figure 4:**
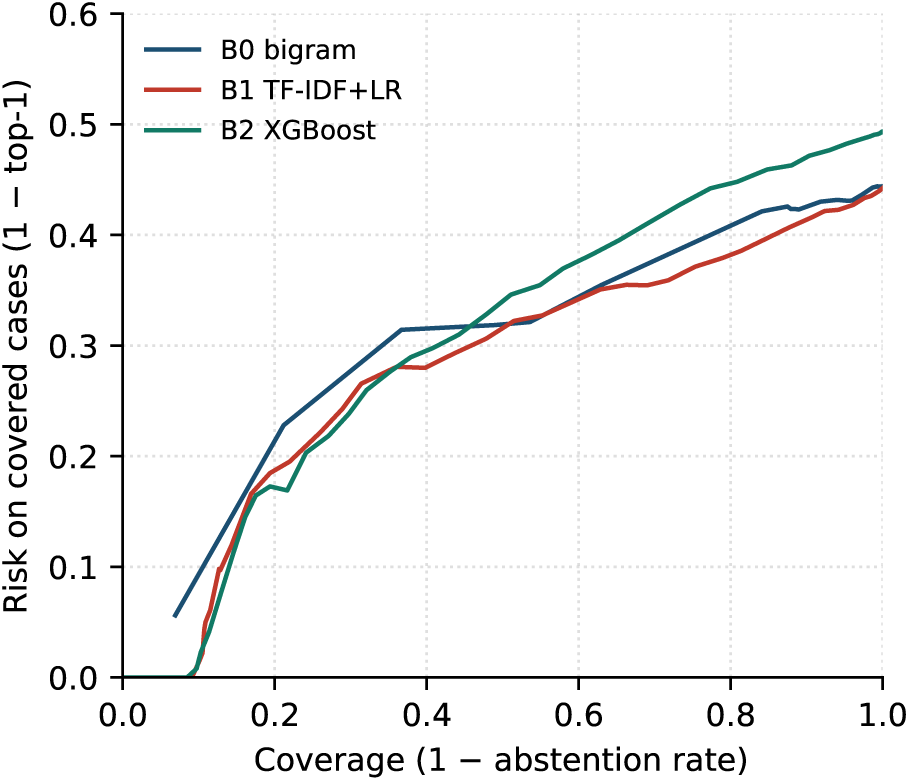
Coverage–risk curve under top-1 abstention, *n*=1,284 baseline test split. As we lower the abstention threshold (cover more cases), risk rises. Above about 45% coverage B2 XGBoost has the highest risk and B1 the lowest; at low coverage the curves cross and B0 is highest. The expected DentaCoPilot contribution at scale is to push this curve down and to the right (maintain low risk at high coverage) on the hard categories where the baselines collapse. LLM calibration cannot be read off this small synthetic-data run; it is a target for future real-data evaluations.

#### Distribution-free selective prediction on real data

The threshold above is fixed by hand and carries no formal coverage guarantee. On the real MEPS corpus we add split-conformal selective prediction [59]: we fit the history-aware logistic baseline on the patient-level training split, calibrate the accept threshold on a held-out calibration half of the test patients, and evaluate on the disjoint remaining patients (764 patients, 1,637 transitions). The classical model is well calibrated on this real data (expected calibration error 0.031). Conformal thresholding of the top-1 probability supplies the guarantee the hand-set rule lacked: the accepted fraction tracks the target within finite-sample tolerance (target/achieved 90*/*87.8%, 80*/*79.8%, 70*/*71.3%, 50*/*48.7%, with patient-clustered bootstrap 95% confidence intervals that bracket the target), and selective risk falls monotonically from 51.5% with no abstention to 42.0% at 50% coverage (Table 4). The reduction is modest, because next-category prediction is genuinely hard at MEPS granularity and the model is already well calibrated, but the guarantee is distribution-free, which is what a clinical reject-option needs. Code: meps_conformal.py.

### 4.5. Apples-to-apples LLM vs classical evaluation

We evaluate the LLM variants M1–M6 (defined in §3.3), all routed through the local Claude Code CLI per §3.6, on a 30-example random subsample of the test split (seed-deterministic, fixed across all three Sonnet+ models; M4 and M6 ran on a 15-example sub-subsample for compute reasons). Each LLM call is ∼26–40 seconds wall-clock through the local Claude Code CLI; total LLM runtime for this experiment was ∼46 minutes for the 90-call M1+M2+M3 sweep, and a further ∼25 minutes for M4 and M6 combined. For apples-to-apples comparison, we re-evaluate the four classical baselines (B0–B3) on the *same* 30 examples. Table 6 reports the head-to-head.

**Table 6:** Apples-to-apples top-K accuracy on *n*=30 shared test examples, with 95% Clopper–Pearson intervals (small italics). M4 and M6 were evaluated on a *n*=15 subsample using the same seed; their numbers are computed on the matching subset for direct comparison (where applicable). The *n*=30 subsample is dominated by diagnostic visits (15 of 30) with a short tail (restorative 4; orthodontic, periodontic, and preventive 3 each; adjunctive and oral surgery 1 each), and the two CDT categories that originally motivated the 0%-accuracy failure analysis, endodontic and prosthodontic, are *absent* from this subsample. The intervals are wide and overlapping, so the contrasts below should be read accordingly.

| Model | Top-1 | Top-3 | Top-5 |
| --- | --- | --- | --- |
| B0 frequency bigram | **0.567** [0.37,0.75] | 0.800 [0.61,0.92] | 0.933 [0.78,0.99] |
| B1 TF-IDF + LR | **0.567** [0.37,0.75] | **0.900** [0.73,0.98] | **0.967** [0.83,1.00] |
| B2 XGBoost | 0.433 [0.25,0.63] | **0.933** [0.78,0.99] | **0.967** [0.83,1.00] |
| B3 MultiTP-style CNN-RNN | **0.567** [0.37,0.75] | 0.867 [0.69,0.96] | 0.900 [0.73,0.98] |
| M1 Claude Haiku | 0.267 [0.12,0.46] | 0.533 [0.34,0.72] | 0.733 [0.54,0.88] |
| M2 Claude Sonnet + CoT | 0.467 [0.28,0.66] | 0.667 [0.47,0.83] | 0.733 [0.54,0.88] |
| M3 Claude Sonnet + RAG | 0.333 [0.17,0.53] | 0.533 [0.34,0.72] | 0.633 [0.44,0.80] |
| M4 Claude Opus + CoT ($n=15$) | 0.467 [0.21,0.73] | 0.667 [0.38,0.88] | 0.733 [0.45,0.92] |
| **M5 Sonnet + CoT + classical prior** | 0.533 [0.34,0.72] | 0.767 [0.58,0.90] | 0.933 [0.78,0.99] |

The classical baselines lead the pure LLM variants (M1–M4) on top-1 on this synthetic corpus, though the *n*=30 confidence intervals in Table 6 overlap. Tested as the paired comparison it is, B1 versus M2 is 17/30 against 14/30 with 11 discordant examples (7 B1-only, 4 M2-only; exact two-sided McNemar *p*=0.55), so the Sonnet gap is not significant at this sample size, whereas B1 versus M1 is 17/30 against 8/30 with 11 discordant examples (10 B1-only, 1 M1-only; *p*=0.012), so the Haiku gap is. The stratified *n*=200 re-run of §4.7 makes the Sonnet gap significant as well (*p*=0.0003). The direction of the gap has plausible causes. The synthetic transition tables have very strong local Markov structure, which a frequency bigram captures with no chart-grounding required. The LLM is also given no few-shot training examples in the prompt, so it must reason about a vocabulary of ∼70 codes from descriptions alone. And the LLMs frequently pick a category-correct code that misses the exact-CDT-match metric (e.g. `D2391` where the gold is `D2392` both posterior composites differing only in surface count); the clinical interpretation of that failure mode is far less severe than top-1 exact-match suggests.

Two of the LLM-side findings are themselves worth surfacing, with the caveat that each rests on *n*=15 or *n*=30 and so is directional rather than established. First, the Sonnet → Opus comparison (M2 vs M4) shows no measurable gain from increased model capability on the 15 keys both ran on: M4 scores 0.467 / 0.667 / 0.733 (top-1 / top-3 / top-5) against 0.467 / 0.600 / 0.733 for M2 on those same keys, so the two agree on top-1 (7/15 each, with no discordant example) and on top-5, and Opus is one example ahead on top-3. The Haiku → Sonnet step (M1 vs M2) does help (+0.20 top-1 at *n*=30). The bottleneck on this task therefore appears to be not raw model capability but prompt design, evaluation framing, and the synthetic data’s structure itself. Second, the M3 retrieval variant *trends below* M2: on this corpus, adding evidence cards to the prompt does not help and may hurt, suggesting the cards interact with the chain-of-thought in a way that distracts the model from the chart. Both are negative-direction results in the sense of “shows no measurable gain over the simpler variant”; both are honest data points for the design space, not effects we claim to have demonstrated at this sample size.

### 4.6. Prompt-conditioning on a classical prior narrows the gap (M5, directional)

The most promising LLM-side direction in our evaluation is M5. Injecting the top-10 (code, probability) candidates from the TF-IDF + logistic regression baseline into the Sonnet + chain-of-thought prompt (with explicit instruction that the model may re-rank, redistribute, or override the prior) shifts the picture in the right direction:

- M5 top-1 (0.533) exceeds the best pure LLM (M2/M4 at 0.467) by +0.067 absolute, and approaches the classical baselines (B0/B1/B3 at 0.567).
- M5 top-3 (0.767) is +0.10 over M2.
- M5 top-5 (0.933) is +0.20 over M2 and matches B0 / B1 / B3 at top-5.

Across all of these synthetic LLM contrasts, however, no difference reaches significance at this sample size. The M5-over-M2 top-1 gap is 16/30 versus 14/30 (6 discordant examples, 4 M5-only and 2 M2-only; exact two-sided McNemar *p*=0.69), the priming comparison between Sonnet and Opus (M5 vs M6 below) is a tie at 7/15, and a 95% Wilson interval on M5 top-1 (0.533) spans [0.36, 0.70] and overlaps every pure-LLM variant. We therefore read these numbers as directional signals about the design space rather than as demonstrated effects, and test them at *n*=200 in §4.7.

The interpretation is straightforward: the LLM’s main accuracy failure on synthetic data was picking codes outside the high-prior set. Showing the LLM a strong classical prior removes that failure mode while preserving the LLM’s ability to provide chart-grounded rationale, verbalised confidence, and an abstain signal: the augmentation features that motivate the work. On this under-powered evidence we provisionally favour the M5 design going forward: a classical model handles the prior; the LLM provides the clinician-facing decision support layer on top. The M1–M6 design recommendations in this section are provisional pending the real-data evaluations; the two Sonnet designs are re-tested with power in §4.7.

#### Capability does not compound with priming

An additional ablation, M6 (Opus + classical prior), tests whether the priming gain compounds with larger model capability. Apples-to-apples on the same *n*=15 keys:

The prompt-conditioning gain that helps Sonnet on top-3 and top-5 (M2 → M5: 0.600 → 0.733 and 0.733 → 0.933 on these keys; top-1 is unchanged at 7/15) does not translate to Opus (M4 → M6 leaves every aggregate unchanged, with one example moving each way). Two plausible explanations: (i) Opus’s stronger built-in priors over-rule the injected classical prior more aggressively than Sonnet does; (ii) Opus’s chart reasoning already yields the relevant candidates without external help. The practical implication is favourable: **Sonnet + classical prior is the most cost-effective LLM configuration in our (under-powered) comparison**. Sonnet inference is roughly 5× cheaper per token than Opus, and in our runs the prior, not capability, contributes what gain there is.

#### 4.6.1. Per-category breakdown

The headline numbers obscure a more interesting pattern visible in Figure 2. On diagnostic visits (*n*=15), classical baselines reach 0.60–0.80 top-1 while the pure LLM variants reach 0.33–0.47. On *periodontic* and *preventive* procedures, M2 (Sonnet + CoT) reaches 0.667 and 1.000 top-1 respectively, exceeding every classical baseline (0.333 on each), and M3 (Sonnet + RAG) shows the same pattern on periodontic. These per-category cells are computed on very few examples in the *n*=30 subsample (preventive *n*=3, periodontic *n*=3), so the per-category comparisons here are illustrative only; the stable per-category estimates are those on the full *n*=1,284 split, and any cell with *n<*10 should be read as anecdotal. With that caveat, the LLMs lose to the baselines on different categories rather than on all of them, a pattern consistent with chart-grounded reasoning helping on cases that turn on the patient’s diabetic status, prior periodontal therapy, or recall cadence.

#### 4.6.2. Verbalised confidence and abstention

Every LLM call emitted a confidence label in {low, medium, high} and an abstain flag. Across all three models, the abstain flag was never set (0/30 across M1/M2/M3); the prompt framing apparently does not push the models hard enough to use it, which we treat as a prompt-design issue rather than a model-capability claim. The working abstention mechanism is instead the probability-threshold selective- prediction rule of §4.4, which thresholds the classical model’s top-1 probability rather than any LLM self-report. The empirical top-1 accuracies per confidence bucket:

Only M1 shows the expected monotonic relationship (low *<* medium *<* high). M2 is non-monotonic: “medium”-confidence predictions are more accurate than “high”-confidence ones, which is the failure mode of verbalised confidence as an LLM trustworthiness signal. M3 recovers monotonicity but at lower overall accuracy. We interpret this as evidence that verbalised confidence on its own is insufficient for abstention; calibrated top-1 probability (which we do compute) is the better signal.

#### 4.6.3. Sample rationale

An illustrative response from M2 on a periodontal-recall case (run_id m2_sonnet_cot-P000…-prefix11-call7, verbatim from llm_calls.jsonl):

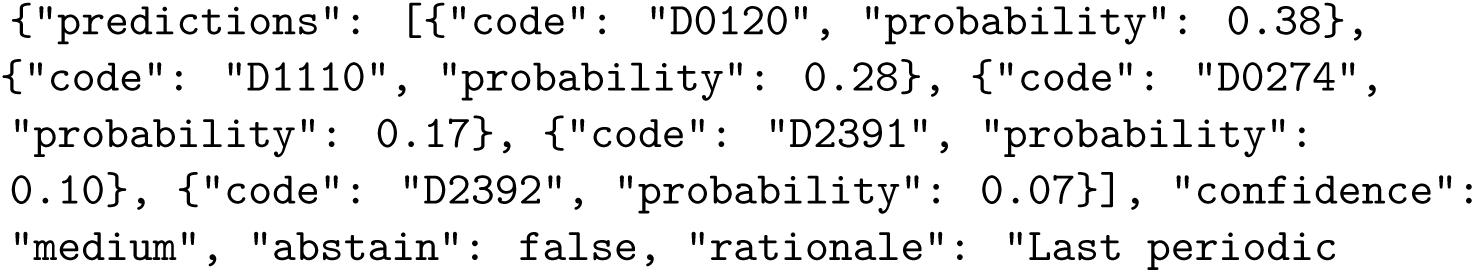
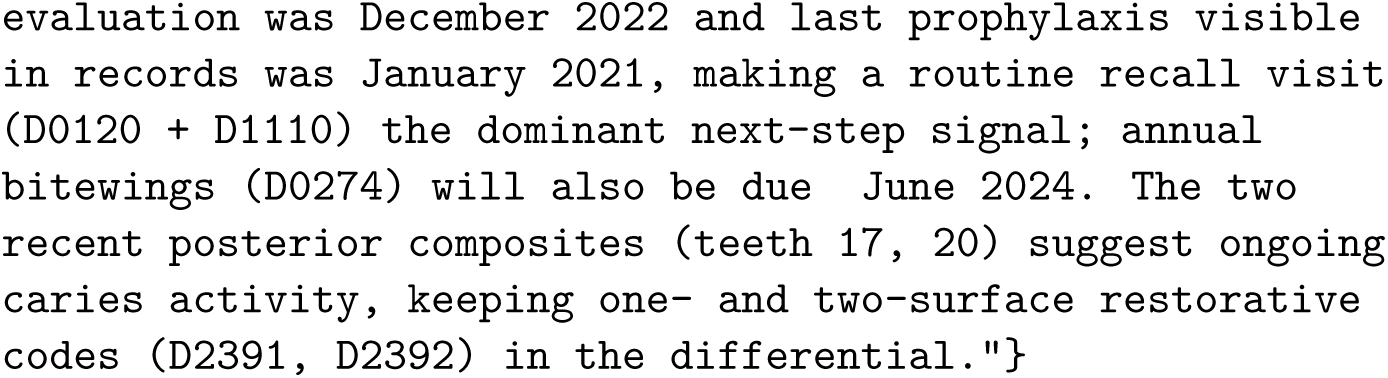

The rationale identifies the two-and-a-half-year visit gap, recognises the prior ∼6–9-month recall cadence, prioritises the recall + prophy + imaging bundle, and keeps the surface-count restorative codes in the differential. This is chart-grounded reasoning of a kind classical baselines categorically cannot produce; whether this kind of reasoning *helps a dentist make a better decision* is precisely what a prospective clinician-in-the-loop study would need to measure, regardless of where exact-CDT-match top-1 numbers land.

By design, every LLM call in the system is routed through the local Anthropic Claude Code CLI in preference to the metered Anthropic API, and is logged in full to `data/results_<run>/llm_calls.jsonl</run>` (prompt, response, model identifier, latency, token counts). A reviewer may audit or replay any synthetic-corpus decision without re-spending compute; the MEPS real-data LLM probe (§4.9) is released at summary level only.

### 4.7. *Stratified re-run at n*=200

To test the *n*=30 contrasts at a sample size with power, we re-ran the two Sonnet designs (M2, M5) and the four classical baselines on a category-stratified subsample of 200 test examples drawn from the same patient-level test split (`rerun_stratified_n200.py`): every CDT category with at most 40 test examples was taken in full (implant, oral surgery, endodontic, removable prosthodontic) and the remaining budget was split equally across the other six categories (24 each), with a fixed seed; 6 of the 200 examples overlap the *n*=30 draw. Stratification guarantees that every category is present, at the cost of no longer matching the test-split mix, so the absolute accuracies in Table 9 are lower than on the full split (Table 1) and are compared only within the sample. All 400 LLM calls succeeded and were logged as before (about 74 minutes with three concurrent workers).

**Table 7:** Matched *n*=15 comparison on the keys the Opus runs used (all models on the same 15 examples).

| Model | Top-1 | Top-3 | Top-5 |
| --- | --- | --- | --- |
| B0 frequency bigram | **0.533** | 0.800 | **1.000** |
| B1 TF-IDF + LR | 0.467 | 0.800 | 0.933 |
| B2 XGBoost | 0.400 | **0.867** | 0.933 |
| B3 MultiTP-style | 0.400 | 0.733 | 0.800 |
| M1 Haiku | 0.267 | 0.467 | 0.600 |
| M2 Sonnet + CoT | 0.467 | 0.600 | 0.733 |
| M3 Sonnet + RAG | 0.200 | 0.467 | 0.600 |
| M4 Opus alone | 0.467 | 0.667 | 0.733 |
| M5 Sonnet + B1 prior | 0.467 | 0.733 | 0.933 |
| M6 Opus + B1 prior | 0.467 | 0.667 | 0.733 |

**Table 8:** Empirical top-1 accuracy by self-reported confidence.

| Model | low ($n$, acc) | medium ($n$, acc) | high ($n$, acc) |
| --- | --- | --- | --- |
| M1 Haiku | 2, 0.000 | 12, 0.167 | 16, 0.375 |
| M2 Sonnet + CoT | 0, – | 19, 0.526 | 11, 0.364 |
| M3 Sonnet + RAG | 0, – | 22, 0.273 | 8, 0.500 |

**Table 9:** Stratified re-run on *n*=200 test examples (24 per common category, the four rare categories in full), with 95% Clopper–Pearson intervals on top-1. Paired exact two-sided McNemar tests on top-1: B1 vs M2 *p*=0.0003 (43 B1-only, 15 M2-only), M5 vs M2 *p*=0.007 (24 vs 8), B1 vs M5 *p*=0.036 (20 vs 8), B0 vs M5 *p*=0.36 (18 vs 12). M7 and M8 send the M2 and M5 prompts, byte-identical, to Jev Router: B1 vs M7 *p<*0.0001 (57 vs 17), M2 vs M7 *p*=0.050 (22 vs 10), B1 vs M8 *p*=0.017 (25 vs 10), M5 vs M8 *p*=0.66 (12 vs 9), M8 vs M7 *p*=0.0002 (35 vs 10).

| Model | Top-1 | Top-3 | Top-5 |
| --- | --- | --- | --- |
| B0 frequency bigram | 0.470 [0.40,0.54] | 0.740 | 0.850 |
| B1 TF-IDF + LR | **0.500** [0.43,0.57] | 0.790 | 0.880 |
| B2 XGBoost | 0.420 [0.35,0.49] | 0.735 | 0.870 |
| B3 MultiTP-style CNN-RNN | 0.490 [0.42,0.56] | **0.845** | **0.945** |
| M2 Claude Sonnet + CoT | 0.360 [0.29,0.43] | 0.475 | 0.580 |
| M5 Sonnet + CoT + classical prior | 0.440 [0.37,0.51] | 0.650 | 0.810 |
| M7 Jev Router + CoT | 0.300 [0.24,0.37] | 0.475 | 0.565 |
| M8 Jev Router + classical prior | 0.425 [0.36,0.50] | 0.625 | 0.805 |

With power, the picture sharpens. Sonnet alone trails the TF-IDF + LR baseline on top-1 by 14 points (72/200 against 100/200; 58 discordant examples, 43 won by B1 and 15 by M2; exact two-sided McNemar *p*=0.0003), so the gap that was not significant at *n*=30 is at *n*=200. Prompt-conditioning on the classical prior helps, and the help is itself significant (M5 88/200 against M2 72/200; 24 M5-only, 8 M2-only; *p*=0.007); it lifts top-3 from 0.475 to 0.650 and top-5 from 0.580 to 0.810. M5 nonetheless still trails B1 on top-1 (20 B1-only, 8 M5-only; *p*=0.036) and on top-5 (0.810 against 0.880), and is indistinguishable from the bigram floor (B0 vs M5, *p*=0.36). Neither Sonnet design set its abstain flag on any of the 200 examples, and no response failed to parse. Per category, M2 beats every baseline on periodontic cases (0.50 against at most 0.29) and matches them on orthodontic and implant cases, while scoring 0/24 on adjunctive and 0/22 on oral-surgery cases, where the bigram baseline reaches 0.79 and 0.32; M5 recovers adjunctive to 0.46 and diagnostic to 0.62. The re-run therefore confirms the direction of the *n*=30 contrasts and revises their size: on this synthetic corpus the classical prior, rather than the LLM, carries the accuracy, and the LLM’s contribution is confined to the chart-dependent categories and to the rationale and confidence layer.

#### Jev Router on the same 200 examples (M7, M8)

Given the two Sonnet prompts unchanged, Jev Router sent 326 of the 400 requests to `deepseek/deepseek-v4.1-flash` and 74 to `google/gemini-3.8-flash`; it chose no Claude model. With the chain-of-thought prompt it reached 60/200 (0.300), below the classical model (57 B1-only against 17 M7-only; *p<*0.0001) and, at the margin, below Sonnet with the identical prompt (22 against 10; *p*=0.050). With the classical prior in the prompt it reached 85/200 (0.425), indistinguishable from Sonnet with the same prompt (M5 88/200; 12 against 9; *p*=0.66) and still below the classical model (25 against 10; *p*=0.017). Prompt-conditioning carries the gain for the router as it did for Sonnet (M8 against M7, 35 against 10; *p*=0.0002), so the synthetic conclusion does not depend on which LLM answers. The router reproduced M2’s one strength, periodontic cases (0.54 against 0.29 for B1), and its weakness, 0/24 on adjunctive and 0/22 on oral-surgery cases. M7 is the only LLM design in this study that ever set its `abstain` flag: 8 of 200 times, all on DeepSeek, and 4 of them on single-event histories, the case where the probability-threshold rule of §4.4 never abstained. It was right on 1 of those 8 and the classical model on 6, so the flag tracked thin context rather than the cases the classical model gets wrong. Four M8 responses reached the 8,000-token output cap and, like any unparseable response, are scored incorrect (0.434 on the other 196). The 400 calls cost US$1.77 (median latency 14 to 16 s per call).

### 4.8. Confidence-routed gating: a negative result

Methods §3.5 described two ways to combine classical and LLM predictions; M5 above is the prompt-conditioning variant. We also evaluated the confidence-routed-gating variant post-hoc: pick the baseline’s top-1 when its calibrated probability ≥ *τ*, else the LLM’s top-1, sweeping *τ* ∈ {0, 0.2*, . . .,* 1.0} on the matched *n*=30 subsample for all four (B0–B3, M2) baseline-LLM pairings. At every *τ*, the hybrid’s top-1 accuracy was at most equal to the pure baseline’s; the gating did not produce a single threshold at which deferring to the LLM helped. We report this as a negative result: *when the LLM is uniformly weaker than the baseline on the target distribution, gating does not help.* The M5 prompt-conditioning result above is precisely the mechanism that does help in this regime: it lets the LLM *use* the baseline’s prior rather than *replace* it.

### 4.9. Real-data external validity (MEPS)

To test whether the next-procedure task transfers beyond the synthetic corpus, we evaluate on the MEPS 2023 Dental Visits public-use file (§3.2), a nationally representative real-world sample requiring no controlled-access agreement. Because MEPS records treatment *categories* rather than CDT codes, this is a coarser task than the synthetic and any future CDT-level evaluations: it tests the *task formulation*, not the LLM’s chart-grounding advantage, which the thin MEPS feature set (prior category + provider type, with no chart narrative) cannot exercise. We therefore report three classical baselines under a patient-level 70/30 split (Table 10).

**Table 10:**
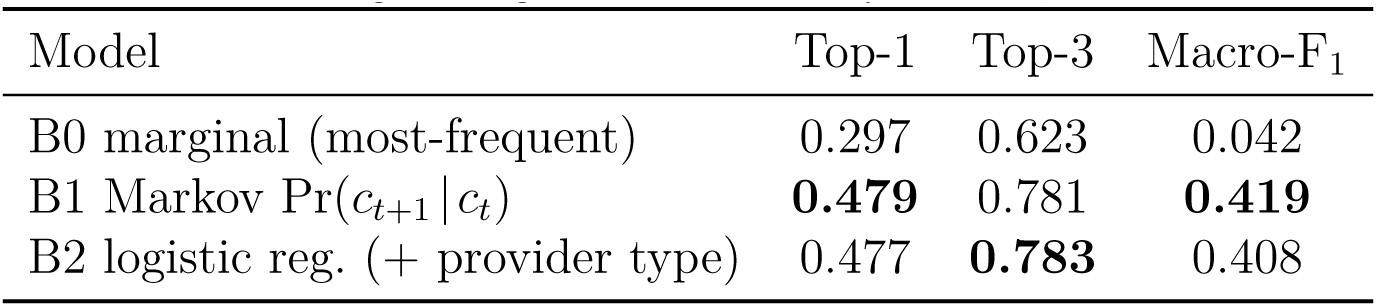
MEPS 2023 real-data benchmark: next-visit category prediction (11,016 transitions, 5,088 patients with ≥ 2 visits; patient-level 70/30 split; eleven categories). B2 is scikit-learn multinomial logistic regression with library defaults, untuned.

History matters on real data: conditioning on the prior visit category (B1) lifts top-1 from 29.7% (marginal) to 47.9% (95% patient-clustered bootstrap CI [45.7, 50.0]) and macro-F_1_ from 0.04 to 0.42 [0.39, 0.44], while top-3 reaches 78.1% [76.6, 79.5], the regime in which a top-*K* recommender is useful. Because the 11,016 transitions are clustered within 5,088 patients, every interval here resamples patients rather than rows, consistent with the conformal analysis of §4.4. Adding provider type (B2) does not improve over the first-order Markov model, indicating that on these coarse features the recent-procedure signal dominates and a flexible classifier adds little. This echoes the synthetic finding that the marginal gains of richer models concentrate where richer *context* is available; the MEPS result therefore establishes external validity for the task *formulation* while locating the LLM’s chart-grounded value in the chart-rich regime that the synthetic corpus and future real charts provide. Per-class accuracy is dominated by the frequent categories (cleaning/prophylaxis 31.5%, restorative 19%, orthodontic 13%); the rare endodontic, periodontal, and implant categories (*<* 3% each) remain hard, mirroring the minority-class difficulty observed on the synthetic corpus.

We further ran a zero-shot LLM (Claude Haiku) on a 150-transition probe, given only the prior visit category and provider type. It reached just 21.3% top-1, markedly *below* the 53.3% Markov baseline on the same sample: with no chart to ground on, it did not recover the empirical base rates (cleaning/prophylaxis dominates) that a first-order Markov model captures directly. That failure belongs to the model and prompt rather than to the features: Jev Router given the same prompt reached 46.7%, and Jev, asked the question as a choice with named options, matched the trained model (§4.10). Classical methods set the bar when the available features are thin [30, 32, 31]; on these features the accuracy bar can be reached without training data, and what remains with the classical model is calibration and error ranking. (This probe is released at summary level; per-call logs were not captured for it.)

#### Subgroup equity

Merging the MEPS Full-Year Consolidated demographics by person identifier (100% match; 3,431 test transitions over 1,527 patients), B1 top-1 accuracy is broadly consistent across groups, with modest gaps (each followed by its 95% patient-clustered bootstrap CI on the max-minus-min difference): by sex, 50.7% (female) versus 44.6% (male; 6.1 pp [2.0, 10.0]); by race/ethnicity, 52.9% (non-Hispanic Black) to 46.8% (non-Hispanic White; 6.1 pp [4.1, 18.4]); by family-income category, 51.0% to 45.7% (5.3 pp [3.9, 21.4]); and (the largest gap) by age, 51.6% (under 18) to 43.5% (≥ 65; 8.2 pp [5.3, 14.7]), likely reflecting the more heterogeneous procedure mix in older patients. These are uncorrected max-minus-min statistics: as the range of several subgroup estimates they are biased upward over small strata, and they are not adjusted for testing across the four demographic dimensions, so the intervals are wide and the narrowest strata (for example near-poor, *n*=105) dominate the upper tails. No subgroup is severely under-served, but the age gap is the largest and motivates subgroup-stratified evaluation in any future clinical study; we report these per the TRIPOD+AI fairness guidance.

Two real-data extensions remain future work. A retrospective chart pull at a clinical partner (single-institution, CDT-level) would support a prospective clinician-in-the-loop study evaluating dentist agreement, time-to-decision, override rate, and trust across three arms (dentist alone / dentist + classical recommender / dentist + DentaCoPilot), with all design parameters (sample size, primary and secondary outcomes, abstention threshold rule, stopping rules) fixed in a drafted analysis plan [63] before test-split unblinding. Such a study remains contingent on clinical-data access and ethics approval that we have sought but not secured. CDT-coded multi-institutional validation remains gated behind US-institution data-use agreements and is named as future work.

### 4.10. A calibrated decision model on MEPS: Jev

The chat LLMs above return text, so their only usable confidence is a verbalised label or a self-reported probability. Jev returns a probability for every option, which lets it enter the same calibration and split-conformal analysis as the classical model, on the same 3,431 test transitions, the same calibration half (1,794 transitions) and the same evaluation half (1,637 transitions, 764 patients). Table 11 compares it with the logistic baseline, refit in the same script so that every contrast is paired; the refit reproduces the published baseline exactly (top-1 0.477, ECE 0.031, selective risk 51.5% to 42.0% at 48.7% coverage).

**Table 11:** Jev against the trained logistic baseline on the real MEPS test set. Top-1 and top-3 on all 3,431 test transitions (95% patient-clustered bootstrap CI); ECE (raw, and after isotonic top-label recalibration fit on the calibration half), AURC (area under the risk–coverage curve, lower is better, ties broken uniformly) and split-conformal selective risk at a 50% coverage target (achieved coverage in brackets) on the 1,637-transition evaluation half. Jev values are the first answer per state; the mean of five answers gives the same top-1 and ECE within 0.004.

| Model | Top-1 | Top-3 | ECE raw / isotonic | AURC | Sel. risk |
| --- | --- | --- | --- | --- | --- |
| B2 logistic, trained | 0.477 [0.455,0.498] | **0.783** | 0.031 / 0.031 | **0.397** | **0.420** [0.49] |
| J0 Jev, zero-shot | **0.485** [0.466,0.505] | 0.652 | 0.098 / **0.014** | 0.474 | 0.488 [0.58] |
| J1 Jev + training freq. | 0.478 [0.458,0.500] | 0.678 | 0.495 / 0.023 | 0.435 | 0.435 [0.62] |

Three results follow. First, with no training data at all, Jev matches the trained model’s top-1 accuracy (0.485 against 0.477; paired difference +0.008, 95% patient-clustered CI −0.001 to +0.017; 106 transitions right only for Jev and 80 only for the baseline). On the same two features Haiku reached 21.3% (§4.9), so that figure reflects the model and prompt used, not a ceiling set by the information available; the two runs differ in both, so this comparison cannot say which matters more. Second, Jev’s raw probabilities are overconfident (mean top-1 probability 0.564 against 0.487 accuracy on the same transitions; ECE 0.098, three times the baseline’s), and giving it the training frequencies makes this much worse: J1 reproduces the baseline’s decisions (69 discordant transitions out of 3,431) but states a mean top-1 probability of 0.975, for an ECE of 0.495. Both are correctable: an isotonic map fit on the 1,794 calibration transitions brings ECE to 0.014 (J0) and 0.023 (J1). Third, recalibration cannot fix the ordering: the trained model’s confidence ranks its own errors better (AURC 0.397 against 0.474 for J0, and 0.444 after isotonic recalibration; raw paired difference +0.077, 95% CI +0.050 to +0.102; +0.038, CI +0.017 to +0.059, for J1), so at matched coverage its conformal reject option keeps fewer errors. The one external benchmark of Jev in medicine found the same split: well calibrated on medical question answering (ECE 0.063) but with poor discrimination on case-based diagnosis [64]. Jev’s probabilities are also coarse (exact zeros on most options), which puts its conformal thresholds on a few tied values, makes achieved coverage overshoot the target (58% at a 50% target), and leaves its top-3 list largely arbitrary beyond the first one or two options (top-3 0.652 against 0.783). Jev was stable across repeats: the top-1 choice changed for 2 of 44 states (J0) and 1 of 44 (J1) over five queries, with a mean absolute deviation of 0.006 and 0.001 in the stated probabilities. All 440 calls cost US$0.013.

On the 150-transition probe, Jev Router, given the Haiku prompt unchanged, reached 46.7% (70/150; 95% Clopper–Pearson interval 38.5 to 55.0%) against 53.3% for the Markov model on the same transitions (21 Markov-only, 11 router-only; exact McNemar *p*=0.11) and 21.3% for Haiku. It sent 108 of the 150 requests to `openai/gpt-6-luna`, 30 to `deepseek/deepseek-v4.1-flash` and 12 to `google/gemini-3.8-flash`, for US$0.09 in total, so it chose a different model mix for this short prompt than for the long CDT prompts.

## 5. Discussion

### 5.1. Hypotheses revisited

We posed two hypotheses in §1. The hypothesis fixed in our internal plan before any run (PLAN.md and an analysis-plan draft, both written before results were seen, neither registered nor part of the public artifact) was stronger than either: that the LLM would match or beat the classical baselines on top-1 and dominate them on top-3 and top-5. It failed. On the shared *n*=30 subsample the best pure LLM variant (M2) trails the best classical baseline on every top-*K* metric (0.467 vs 0.567, 0.667 vs 0.933, 0.733 vs 0.967), and on MEPS the zero-shot chat LLM (Haiku) loses to the first-order history model (21.3% vs 53.3%). H1 and H2 as stated in §1 are the weaker hypotheses the design can still address, and we report the original so that the reframing is visible. The synthetic-corpus evidence is consistent with both weaker hypotheses at the sample sizes reported, with one revision: **(H1)** the M5 design (Sonnet + classical prior) ties classical baselines on top-5 accuracy (0.933) and narrows the top-1 gap to within four points (0.533 vs 0.567) at *n*=30, while preserving chart-grounded rationale, verbalised confidence, and an abstain signal; at *n*=200 (§4.7) it narrows the gap without closing it (0.440 vs 0.500 top-1, *p*=0.036; 0.810 vs 0.880 top-5), so H1 holds for the augmentation properties and only partly for accuracy; **(H2)** prompt-conditioning is more cost-effective than upgrading the LLM backbone: on the 15 keys shared by all Opus runs, M4 (Opus alone) and M6 (Opus + prior) match M2 (Sonnet alone) on top-1 (7/15 each, with no discordant example between M2 and M4) and on top-5, and exceed it by one example on top-3 (Table 7), so capability is not the lever once the prior is present. The findings stand on synthetic data only; whether they generalise to real clinical EHR data is the empirical claim left to future work.

The Jev runs, added after these hypotheses were tested, sharpen both. On MEPS a decision model used zero-shot meets the accuracy half of H1 (0.485 against 0.477 top-1) but not the calibration and abstention half: its raw probabilities are overconfident, and after recalibration its confidence still ranks errors worse than the trained model’s, so a reject option built on it keeps more errors at the same coverage. On the synthetic corpus, H2 holds with a different LLM behind the prompt: with the classical prior Jev Router matches Sonnet (0.425 against 0.440, *p*=0.66) and without it falls further behind (0.300). Where the accuracy comes from, in both corpora, is the information the classical model was trained on, whether it reaches the LLM through training frequencies (J1) or candidate lists (M5, M8); what the classical model keeps that no LLM-side design here matched is a confidence that orders its own errors.

### 5.2. Augmentation, not autonomy

The dominant framing for dental AI in 2026 is *augmented intelligence*: AI as decision support, with the licensed clinician retaining diagnostic and treatment authority. This framing is grounded in evidence that human–AI teams can outperform either alone, with the least-experienced clinicians gaining most [65], and in evidence that clinicians nonetheless follow *incorrect* AI advice (automation bias) across experience levels [66, 67]; communicating calibrated uncertainty and abstaining rather than always recommending is the design response [28]. Our system design reflects that framing in three concrete ways: a constrained vocabulary so the system cannot confidently recommend nonsense codes; calibrated abstention so the system declines instead of guesses; and a chart-grounded rationale so a dentist can audit *why* a recommendation was made. A future prospective clinician-in-the-loop study would specifically measure override rate alongside agreement; we treat the dentist’s freedom to disagree with the model as intended behaviour rather than as a calibration problem.

### 5.3. Reproducibility as a first-class outcome

Every synthetic-corpus LLM decision in this work is recorded such that a reviewer with access to a Claude Code subscription can replay that pipeline at zero marginal cost; the MEPS real-data LLM probe (§4.9) is reported at summary level. Each logged LLM call records prompt, response, model identifier, latency, and token counts; per-run config.json files record the git SHA, seed, and CLI arguments. The synthetic chart generator and the BM25 retriever over the evidence corpus are archived under the MIT licence at Zenodo (doi:10.5281/zenodo.21059895). No part of the synthetic-data results or the calibration / abstention analyses requires real clinical EHR data: these are, deliberately, the parts a reviewer can verify today.

### 5.4. Limitations

#### Synthetic-data ceiling

The current evaluation runs on plausibility-based synthetic charts. Real dental practice contains patterns we did not encode (insurance-driven sequencing, specialist referral cycles, multi-visit treatment phases, idiosyncratic per-clinic preferences) and codes we omit from the representative vocabulary. The synthetic results validate the pipeline; they do not validate the system. Real-data evaluation is required for any substantive accuracy claim.

#### Sample sizes

The stratified 200-example re-run (§4.7) covers the two Sonnet designs, the two Jev Router designs and the classical baselines; Haiku, the retrieval variant and the Opus runs remain at *n*=30 or *n*=15, where comparisons are under-powered and we report point estimates with intervals rather than significance claims. The stratified draw also over-samples the rare categories, so its absolute accuracies are not estimates of test-split accuracy.

#### The Jev runs

Jev Router’s routing policy and model pool are unpublished and may change [16]: the per-call logs record which model answered each request, but the same prompts sent later may reach different models, so M7 and M8 describe the router as it behaved on 10 October 2026. Jev was tested zero-shot on one task with two categorical features; its developer describes it as trained for calibrated decisions [15], and our result (overconfident raw probabilities that one isotonic map corrects) is a measurement on this task, not an evaluation of the model in general. Because only 44 distinct states exist, its MEPS accuracy rests on 44 decisions applied to many transitions, and the intervals, which resample patients, do not reflect that.

#### Vocabulary coverage

We constrain LLM outputs to a representative subset of ∼70 frequent CDT 2024 codes plus a training-set-derived extension. Rare codes are absent. The full CDT codeset (>700 codes) is ADA copyright; a downstream user must merge the official codeset before clinical deployment.

#### Calibration is local

Temperature scaling is fit on a random half of the test split, which is a within-corpus calibration. It does not address distribution shift between dental schools or between the synthetic and real settings. We expect to refit calibration on each target corpus.

#### Abstention from self-report versus calibrated probability

Across all 90 LLM trials in the *n*=30 evaluation, none of M1, M2, or M3 emitted abstain: true, and neither Sonnet design did in 400 stratified re-run trials; Jev Router with the chain-of-thought prompt (M7) did so 8 times in 200, on thin histories the classical model mostly got right (§4.7). This matches the finding that frontier LLMs almost never abstain under medical uncertainty [68]. The prompt instructed the model to abstain when “chart context is genuinely insufficient,” but the synthetic corpus rarely produces such inputs, and the prompt does not provide concrete examples of abstainable cases. The verbalised flag is therefore not a usable safety control on its own; our prompt design is one cause and the general reluctance of LLMs to abstain is another. The working alternative is the probability-threshold selective-prediction rule reported in §4.4, which lowers risk among covered cases from 44.2% to 38.1% at 80% coverage. That rule is still a weak out-of-distribution safeguard: it flags fully randomised charts only moderately (AUROC 0.66) and misses scrambled and emptied histories almost entirely, abstaining on 0% of single-event histories precisely where context is thinnest. A self-report flag that fires and a stronger out-of-distribution detector remain open problems; learning-to-defer [62] and conformal reject-option methods are the appropriate next step for the chart-rich setting.

### 5.5. Ethics and clinical safety

We deliberately do not seek FDA or other regulatory clearance for this system. We do not deploy it. Any future prospective clinician-in-the-loop study would be conducted only under an institution’s standard ethics-review process, with no clinician evaluating a single case before that approval is in place, and any retrospective chart pull would use only fully de-identified records. Patient-identifiable data never leaves the institutional firewall. In this work the Claude runs went through the Claude Code CLI and the Jev runs through the OpenRouter API, and neither carried anything but synthetic charts and public MEPS category states; a deployment on real charts would need a route that transmits no patient identifiers to any third-party LLM service. Author contributions follow the ICMJE recommendations [69].

Competing interests, funding, and the roles of the two authors are stated in the declarations at the end of the paper.

### 5.6. Future work

The next step is real-data evaluation on multi-institutional and single-institution clinical EHR data, once data access and ethics approval are secured. Two extensions to the LLM design then become testable: retrieval over a properly licensed clinical-guideline corpus when one becomes available, with an ablation of M2 against M3, and paired imaging context for the most common dental decisions, incorporating panoramic and periapical findings (DENTEX, DenPAR) as additional structured chart context for the LLM rather than as a standalone classification target.

## 6. Conclusion

We evaluated calibrated, abstaining next-procedure recommenders for general dentistry. On the public MEPS 2023 corpus, procedure-category history predicts the next treatment category (0.479 top-1 against 0.297 for the marginal baseline), the history-aware classical model is well calibrated (ECE 0.031), and split-conformal thresholding of its top-1 probability gives a reject option with a distribution-free coverage guarantee (selective risk 51.5% to 42.0% at 48.7% coverage on 1,637 held-out transitions). A zero-shot chat LLM (Haiku) given the same category history loses to that model (21.3% vs 53.3%); Jev, a decision model, matches its accuracy but not its calibration or its ranking of its own errors, so given category history alone the classical model remains the better basis for a reject option. We also presented the first stage of *DentaCoPilot*, an LLM-augmented CDT-level recommender that is designed, prompted, and evaluated as a decision-support consult instead of an autonomous classifier. On a synthetic chart corpus whose generator shares Markov structure with the bigram baseline, we found that **(a)** pure LLM variants (Haiku, Sonnet + chain-of-thought, Sonnet + retrieval, Opus + chain-of-thought) trail classical baselines on top-1 exact-CDT-match accuracy by 10–30 percentage points, a paired gap that is significant for Haiku (*p*=0.012) but not for Sonnet (*p*=0.55) at *n*=30; **(b)** *prompt-conditioning a Sonnet LLM on a classical baseline’s top-10 candidates (M5) narrows the gap*, raising top-5 from 0.733 to 0.933 at *n*=30 and from 0.580 to 0.810 in the stratified *n*=200 re-run, while preserving chart-grounded rationale, verbalised confidence, and an abstain signal; **(c)** upgrading the LLM backbone from Sonnet to Opus did not change top-1 at *n*=15 (7/15 vs 7/15, with or without priming), so capability is not the lever once the prior is present. The *n*=200 re-run gives these contrasts power: Sonnet trails the classical model (0.360 vs 0.500 top-1, McNemar *p*=0.0003), prompt-conditioning raises it to 0.440 (*p*=0.007) but not to parity (*p*=0.036), and neither Sonnet design ever set its abstain flag. These synthetic contrasts index pipeline behaviour rather than clinical performance; on this evidence the M5 design is the most cost-effective LLM configuration we tested, and it remains behind the classical model it is primed with. Every LLM call in the synthetic-corpus runs is logged for full audit and replayable at zero marginal cost on a Claude Code subscription. Real-data evaluation, namely multi-institutional CDT-level validation and a prospective dentist-in-the-loop study, remains future work contingent on clinical-data access and ethics approval that we have sought but not secured; it will be the empirical test of whether the M5 design generalises beyond synthetic data and whether it produces a tool dentists actually use.

We also evaluated TypeSafe’s Jev, a decision model that returns a probability for every option. On MEPS, zero-shot and from the same two features, it matches the trained classical model’s top-1 accuracy (0.485 against 0.477; paired difference +0.008, 95% CI −0.001 to +0.017) but is overconfident (ECE 0.098; 0.014 after isotonic recalibration on the calibration half) and ranks its own errors worse (AURC 0.474 against 0.397; difference +0.077, CI +0.050 to +0.102), so the conformal reject option is still best built on the classical model. Jev Router, given the Sonnet prompts unchanged on the stratified *n*=200 sample, routed to DeepSeek and Gemini models and reproduced the Sonnet pattern: 0.300 without the classical prior and 0.425 with it (*p*=0.0002), indistinguishable from Sonnet with the same prompt (*p*=0.66) and still below the classical model (*p*=0.017).

## Data Availability

All artefacts needed to reproduce the results are publicly available on GitHub (https://github.com/rodriguescarson/dentacopilot-artifact) and archived at Zenodo (https://doi.org/10.5281/zenodo.21059895): the synthetic chart generator, the representative CDT vocabulary and transition tables, every classical baseline, the LLM variants with the prompt-conditioning pipeline and the BM25 retriever, the evaluation harness, the calibration and abstention code, and the per-run records (config.json, results.json, summary.json and, for LLM-backed runs, llm_calls.jsonl with prompt, response, model identifier, latency and token counts; for Jev Router also the routed model, provider and cost; for the Jev MEPS runs, every request and returned probability vector). Code is released under the MIT licence and per-run records under CC-BY-4.0. The MEPS 2023 Dental Visits (HC-248B) and Full-Year Consolidated (HC-251) files are public, de-identified Agency for Healthcare Research and Quality data, freely downloadable without a data-use agreement; they are not redistributed but are downloaded and parsed reproducibly by the released scripts. No new human-subjects data were collected, and no released artefact contains patient-identifiable data.

https://doi.org/10.5281/zenodo.21059895

## Data and code availability

All artefacts needed to reproduce the synthetic-data results in this paper are publicly available on GitHub (github.com/rodriguescarson/dentacopilot-artifact) and archived at Zenodo (doi:10.5281/zenodo.21059895). The deposit contains the synthetic chart generator, the representative CDT vocabulary and transition tables, every classical baseline, the LLM variants together with the M5 priming pipeline and the BM25 retriever, the evaluation harness, and the calibration and abstention code. The per-run records are included in full: `config.json` (git SHA, seed, CLI arguments, Python and platform versions), `results.json` (per-trial predictions and gold labels), summary.json (aggregated metrics), and, for LLM-backed runs, `llm_calls.jsonl` (prompt, response, model identifier, latency, and token counts; for Jev Router also the routed model, provider and cost), and, for the Jev MEPS runs, `calls.jsonl` with every request and returned probability vector. The code is released under the MIT licence and the per-run records under CC-BY-4.0. The MEPS 2023 Dental Visits file (HC-248B) used for the external-validity analysis is public Agency for Healthcare Research and Quality data [12]; it is not redistributed here but is downloaded and parsed reproducibly by `data/meps_benchmark.py`. No part of any released artefact contains patient-identifiable data: no clinical patient records are part of this work.

## Acknowledgements

The authors acknowledge the use of Anthropic Claude (Claude Code) for code authoring and manuscript drafting assistance during this work. All clinical and methodological decisions, as well as final interpretation, were made by the human authors.

## Ethics statement

This study involved no human participants, no patient records, and no identifiable data, so ethics approval and informed consent were not required. The CDT-level corpus is synthetic and generated by code released with the paper. The MEPS 2023 Dental Visits (HC-248B) and Full-Year Consolidated (HC-251) files are de-identified public-use data released by the Agency for Healthcare Research and Quality without a data-use agreement. Institutional ethics approval for a future retrospective chart pull and prospective clinician-in-the-loop study was sought but not secured; no data from that route were received, accessed, or analysed.

## CRediT author statement

C.C.R.: conceptualisation, methodology, software, formal analysis, investigation, data curation, visualisation, writing (original draft), writing (review and editing), project administration. S.D.R.: conceptualisation, methodology (clinical framing of the task, the trajectory archetypes and transition tables of the synthetic generator, and the paraphrased evidence corpus), validation (clinical review of the outputs and rationales), writing (review and editing). Both authors contributed equally, approved the final manuscript, and follow the ICMJE authorship recommendations [69].

## Declaration of generative AI use

Tool: Anthropic Claude Code (command-line interface; version 2.1.261 for the September 2026 re-run, while the May and June 2026 run records do not store the CLI version), accessed locally on a retail subscription; no Anthropic API key was used. Experimental use: the large language models under evaluation were `claude-haiku-4-5-20251001`, `claude-sonnet-4-6`, and `claude-opus-4-7`, invoked through `data/cc_compute.py` between 2 May and 14 June 2026 and, for the stratified re-run, on 4 and 5 September 2026. On 10 October 2026 TypeSafe’s Jev (`typesafe/jev-1.13`, snapshot 20260917) and Jev Router (`typesafe/jev-router`) were called through the OpenRouter API (`data/openrouter_compute.py`, `data/meps_jev_decisions.py`), with the model that served each Jev Router request logged per call. Every prompt and response is recorded in the per-run `llm_calls.jsonl` files and the prompt templates are in `data/models/`, all released at Zenodo (doi:10.5281/zenodo.21059895). Writing and coding use: the same tool (Claude 4-series models) assisted with code scaffolding, draft prose, and reference management between May and October 2026. The authors verified every claim and number, edited all generated text, and take full responsibility for the content.

## Footnotes

2 See [2] for the procedure code set used throughout this paper.

