## Supplementary Material for "A Zero-Shot Decision Model (Jev) Matches a Trained Classifier but Ranks Its Own Errors Worse: Calibrated, Split-Conformal Next-Procedure Recommendation in General Dentistry"

**Version:** rebuilt 2026-10-10 from the master (`draft/paper.tex`), after the Jev runs were added.

This supplement maps the manuscript against **TRIPOD+AI** (Collins et al., *BMJ* 2024; `10.1136/bmj-2023-078378`) and **TRIPOD-LLM** (Gallifant et al., *Nature Medicine* 2025; `10.1038/s41591-024-03425-5`). DECIDE-AI (Vasey et al., *Nat Med* 2022) applies to any future prospective clinician-in-the-loop study; CLAIM applies only to a future imaging-grounding arm.

**Stage note.** This is a development + proof-of-concept paper evaluated on (i) a synthetic chart corpus and (ii) a public, de-identified, aggregate real-world benchmark (MEPS). It involves **no human-subjects data and no clinical deployment**; clinical-validation, fairness-by-sociodemographic-group, and patient-involvement items are therefore reported as *N/A at this stage* or *Planned (future prospective study)*. Items follow the numbering of the official TRIPOD+AI checklist (Collins et al. 2024, Table 1) and the domain structure of TRIPOD-LLM.

Status key: **Y** addressed · **P** partial · **NA** not applicable at this stage · **GAP** acknowledged limitation, not addressed.

---

### TRIPOD+AI checklist (27 items)

| # | Item | Status | Location / note |
|---|---|---|---|
| 1 | Title identifies model development/evaluation, population, outcome | Y | Title states the finding (a zero-shot decision model, Jev, matches a trained classifier but ranks its own errors worse) and names the task (next-procedure recommendation), the evaluation objects (calibration, split-conformal abstention) and the population (general dentistry). |
| 2 | Abstract per TRIPOD+AI-for-Abstracts | P | Unstructured abstract (journal style). It reports data sources, sample sizes, primary metrics with 95% intervals, calibration (ECE 0.031), conformal abstention (selective risk 51.5% to 42.0% at 48.7% coverage) and the paired test results, so the TRIPOD+AI-for-Abstracts content items are covered without headings. |
| 3a | Healthcare context & rationale | Y | §Introduction (diagnostic-only commercial market; the "what next" gap). |
| 3b | Target population, intended use & users | Y | §Introduction (general dentists as users; augmentation, not autonomy). |
| 3c | Known health inequalities across sociodemographic groups | P | Subgroup performance now reported on MEPS by sex/age/race-ethnicity/poverty (§Real-data, "Subgroup equity"); disparities characterised. |
| 4 | Study objectives (development / evaluation / both) | Y | §Introduction (H1, H2; develop + evaluate). |
| 5a | Data sources (development & evaluation) | Y | §Datasets (synthetic corpus; MEPS 2023 HC-248B). |
| 5b | Dates of data collection | P | MEPS = calendar year 2023; synthetic spans a simulated 5-yr timeline (generated, not collected). |
| 6a | Study setting, centres, locations | NA | Synthetic; MEPS = US nationally representative sample (no centre structure). |
| 6b | Eligibility criteria | Y | MEPS: persons with 2 or more dental visits; synthetic: all generated charts (§Datasets). |
| 6c | Treatments received & how handled | Y | Procedures/CDT codes are the prediction target (§Task formulation). |

| # | Item | Status | Location / note |
|---|---|---|---|
| 7 | Data preparation & quality checks | Y | §Methods (CDT-2024 constrained vocabulary; MEPS flag→primary-category mapping; seeds). |
| 8a | Outcome definition, horizon, assessment | Y | §Task formulation (next procedure = next CDT code; next visit category for MEPS). |
| 8b | Subjective outcome assessment | NA | Outcome = recorded procedure code (objective). |
| 8c | Blinding of outcome assessment | NA | As above. |
| 9a | Predictor selection rationale | Y | Chart history features (§Models: bag-of-codes, TF-IDF, sequence). |
| 9b | Predictor definitions, timing, blinding | Y | §Task/§Models (prior procedures + context, measured before the predicted event). |
| 9c | Subjective predictor assessment | NA | Predictors are recorded codes (objective). |
| 10 | Sample size & justification | P | Synthetic n=500 charts (8,427 examples: 5,878 train / 1,265 dev / 1,284 test); MEPS 11,016 transitions / 5,088 patients; **LLM arm n=30 is explicitly acknowledged as underpowered** and paired exact McNemar tests with discordant counts are reported. |
| 11 | Missing data handling | P | Synthetic complete by construction; MEPS uses available treatment flags (missing/refusal codes excluded). |
| 12a | Data use / partitioning | Y | Patient-level 70/15/15 (synthetic); patient-level 70/30 (MEPS). |
| 12b | Predictor handling / transformations | Y | §Models (TF-IDF weighting, one-hot, XGBoost features). |
| 12c | Model type, building, hyperparameters, internal validation | Y | §Models (B0–B3, M1–M6, Jev designs M7, M8, J0, J1; constrained decoding; temperature scaling; isotonic recalibration for Jev on MEPS). |
| 12d | Heterogeneity across clusters | NA | No multi-cluster estimation. |
| 12e | Performance measures (discrimination, calibration, utility) | Y | top-1/3/5, macro-F1; calibration (temp scaling) + coverage–risk (§Calibration/abstention). |
| 12f | Model updating | NA | No updating; calibration is reported, not iterative updating. |
| 12g | How predictions computed (formula/code) | Y | Open-source code; every LLM call logged (`llm_calls.jsonl`). |
| 13 | Class-imbalance methods | P | No resampling; minority-class (endodontic/implant/perio) difficulty reported honestly. |
| 14 | Fairness approaches | Y | B1 top-1 accuracy reported by sex, age band, race/ethnicity, and family-income category with max-min gaps (largest = age, 8.2 pp); §Real-data, "Subgroup equity". |
| 15 | Model output type & thresholds | Y | top-K probability distribution + verbalised confidence + explicit abstain (§Task). |
| 16 | Development vs evaluation differences | Y | Synthetic (CDT-level) vs MEPS (category-level) differences stated explicitly (§Datasets, §Real-data). |
| 17 | Ethical approval / consent | Y | Ethics statement in the Declarations: no human participants, no patient records, public de-identified MEPS data; approval and consent not required. Approval for a future retrospective chart pull and prospective clinician study was sought but not secured; no such data were accessed. |
| 18a | Funding & funder role | Y | Funding statement in the Declarations: no specific grant; compute on a retail Claude Code subscription and OpenRouter usage paid by Celabe; no funder role. |
| 18b | Conflicts of interest | Y | Competing-interests declaration in the Declarations (no vendor relationships; C.C.R. founder of Celabe; no patent or product). |

| # | Item | Status | Location / note |
|---|---|---|---|
| 18c | Protocol accessibility | P | Methods fully documented; the internal analysis plan fixed before the runs (PLAN.md) is described in §Discussion (Hypotheses revisited), including the pre-specified hypothesis that failed; a protocol for any future prospective study would be registered at that time. |
| 18d | Registration | P | This stage was not registered (the manuscript says so; nothing is described as pre-registered); any future prospective clinical study would be registered before data collection. |
| 18e | Data availability | Y | Synthetic generator open-source; MEPS public (AHRQ); charting datasets released. |
| 18f | Code availability | Y | All code MIT-licensed; per-call audit logs released. |
| 19 | Patient & public involvement | NA | None at this (pre-clinical) stage; clinicians would be engaged in any future prospective study. |
| 20a | Participant flow | NA/P | Report transition/example counts (no participant-flow diagram; synthetic + aggregate). |
| 20b | Characteristics by source/group | P | MEPS category distribution + synthetic archetypes reported; no sociodemographic breakdown. |
| 20c | Distribution comparison (dev vs eval) | Y | **Generator face-validity check vs MEPS** (marginal r=0.19, transition r=0.50, weak agreement; synthetic accuracy is therefore treated as a pipeline test, not real-world performance); §Datasets. |
| 21 | N participants/events per analysis phase | Y | Test-example/transition counts reported per experiment. |
| 22 | Full model specification | Y | Open-source models + prompts + constrained vocabulary enable reproduction. |
| 23a | Performance estimates with confidence intervals & subgroups | Y | 95% patient-clustered bootstrap CIs (B=1000) on all MEPS metrics (top-1 47.9% [45.7, 50.0], top-3 78.1% [76.6, 79.5], macro-F1 0.42 [0.39, 0.44]; `data/results_meps_2023/ci_fairness.json`) + per-category and per-subgroup breakdowns; §Real-data. Clopper-Pearson intervals on every n=30 cell. |
| 23b | Heterogeneity in performance | NA | No cluster-level performance analysis. |
| 24 | Model-updating results | NA | No updating performed. |
| 25 | Interpretation (incl. fairness) vs objectives & prior work | Y | §Discussion (hypotheses revisited, including the failed pre-specified hypothesis; augmentation framing); subgroup gaps interpreted in §Real-data. |
| 26 | Limitations (representativeness, overfitting, missing data) | Y | §Limitations (synthetic data + circularity caveat; n=30; abstention limits; WEIRD/coarse-MEPS). |
| 27a | Handling poor/unavailable predictors at implementation | Y | Abstention design declines on insufficient context (with honest limits, §Calibration/abstention). |
| 27b | User interaction & expertise required | Y | §Augmentation (licensed dentist retains authority; rationale supports audit). |
| 27c | Next research steps / generalizability | Y | §Future work (a future prospective clinical study; CDT-level multi-institutional validation). |

**TRIPOD+AI summary:** Addressed (Y) ~20 · Partial (P) ~7 · N/A 8 · **Gaps 0.** The three previously-flagged gaps are now closed on the MEPS real-data benchmark: **23a** (95% bootstrap CIs added), **14** (fairness/subgroup accuracy by sex/age/race-ethnicity/poverty with reported gaps), and **3c** (sociodemographic disparities characterised). Remaining N/A items are inherent to a pre-clinical, no-human-subjects stage and are scoped to any future prospective clinical study.

### TRIPOD-LLM (Gallifant et al. 2025): domain-level mapping

TRIPOD-LLM is modular for studies developing/evaluating LLMs in health. Domain-level status (complete exact items via the official fillable tool before submission):

| Domain | Status | Location / note |
|---|---|---|
| Title & abstract (LLM identified) | Y | DentaCoPilot identified as an LLM-based recommender. |
| Background, task, intended use | Y | §Introduction. |
| Data & task definition | Y | §Datasets, §Task formulation. |
| **LLM details: models, versions, access** | Y | Claude Haiku/Sonnet/Opus via Claude Code; TypeSafe Jev (typesafe/jev-1.13, snapshot 20260917) and Jev Router (typesafe/jev-router) via the OpenRouter API on 10 October 2026; versions logged per call, and for Jev Router the model that served each request. |
| **Prompt / instructions reported** | Y | Prompt templates in §Models and in the released code (`data/models/`); every prompt and response in `llm_calls.jsonl`; the Declaration of generative AI use lists tool, model versions, dates, access method and prompt locations. |
| Stochasticity / determinism | P | Temperature/calibration discussed; per-call logs capture variability; quantify run-to-run variance explicitly. |
| Evaluation metrics & comparators | Y | Classical baselines (B0–B3) vs LLM variants; top-K, macro-F1, calibration, coverage–risk. |
| Human oversight / role of clinician | Y | §Augmentation: explicit “augment, not replace”; abstain + rationale for auditability. |
| Uncertainty / abstention | Y | §Calibration & abstention (incl. the honest OOD/over-confidence finding). |
| Fairness / bias | Y | Subgroup accuracy by sex/age/race-ethnicity/poverty on MEPS (max-min gaps 5.3-8.2 pp); §Real-data. |
| Reproducibility / open science | Y | MIT code, full per-call audit logs, public MEPS data, synthetic generator released. |
| Ethics / data governance | Y/NA | No human subjects this stage; §Ethics; any future prospective arm would be under institutional ethics approval. |

**Net:** the manuscript satisfies the LLM-specific transparency, prompt-reporting, reproducibility, and human-oversight items that TRIPOD-LLM emphasises, and the previously-outstanding **fairness/subgroup** and **confidence-interval** items are now addressed on the MEPS real-data benchmark. The remaining items requiring real clinical deployment (prospective performance, clinician workflow/usability) are scoped to a future prospective DECIDE-AI study.
